# Enoxaparin for thromboprophylaxis in hospitalized COVID-19 patients: comparison of 40 mg o.d. *vs* 40 mg b.i.d. The X-COVID19 Randomized Clinical Trial

**DOI:** 10.1101/2021.11.17.21266488

**Authors:** Nuccia Morici, GianMarco Podda, Simone Birocchi, Luca Bonacchini, Marco Merli, Michele Trezzi, Gianluca Massaini, Marco Agostinis, Giulia Carioti, Francesco Saverio Serino, Gianluca Gazzaniga, Daniela Barberis, Laura Antolini, Maria Grazia Valsecchi, Marco Cattaneo

**Affiliations:** Intensive Coronary Care Unit, ASST Grande Ospedale Metropolitano Niguarda, Milan, Italy; Unità di Medicina 2, ASST Santi Paolo e Carlo, Milan, Italy; Dipartimento di Scienze della Salute, Università Degli Studi di Milano, Milan, Italy; Emergency Department, ASST Grande Ospedale Metropolitano Niguarda, Milan, Italy; Divisione di Malattie Infettive, ASST Grande Ospedale Metropolitano Niguarda, Milan, Italy; Struttura Operativa Complessa (SOC) Malattie Infettive II, AUSL Toscana Centro, Ospedale San Jacopo, Pistoia, Italy; Struttura Operativa Semplice (SOS) Chirurgia vascolare, AUSL Toscana Centro, Ospedale San Jacopo, Pistoia, Italy; Emergency Department, Ospedale San Carlo Borromeo, ASST Santi Paolo e Carlo, Milano, Italy; ASL 4 Veneto, Covid Hospital, Jesolo, Italy; Postgraduate School of Clinical Pharmacology and Toxicology, Department of Oncology and Hemato-Oncology, University of Milan, Milan, Italy; Center of Bioinformatics, Biostatistics and Bioimaging, School of Medicine and Surgery, University of Milano Bicocca, Monza, Italy

**Keywords:** pulmonary embolism, thrombosis, COVID-19, enoxaparin

## Abstract

It is uncertain whether higher doses of anticoagulants than recommended for thromboprophylaxis are necessary in COVID-19 patients hospitalized in general wards. This is a multicentre, open-label, randomized trial performed in 9 Italian centres, comparing 40 mg b.i.d. vs 40 mg o.d. enoxaparin in COVID-19 patients, between April 30, 2020 and April 25, 2021. Primary efficacy outcome was in-hospital incidence of venous thromboembolism (VTE): asymptomatic or symptomatic proximal deep vein thrombosis (DVT) diagnosed by serial compression ultrasonography (CUS), and/or symptomatic pulmonary embolism (PE) diagnosed by computed tomography angiography (CTA). Secondary endpoints included each individual component of the primary efficacy outcome and a composite of death, VTE, mechanical ventilation, stroke, myocardial infarction, admission to ICU. Safety outcomes included major bleeding. The study was interrupted prematurely due to slow recruitment. We included 183 (96%) of the 189 enrolled patients in the primary analysis (91 in b.i.d., 92 in o.d.). Primary efficacy outcome occurred in 6 patients (6·5%, 0 DVT, 6 PE) in the o.d. group and 0 in the b.id. group (ARR 6·5, 95% CI, 1·5-11·6). Absence of concomitant DVT and imaging characteristics suggest that most pulmonary artery occlusions were actually caused by local thrombi rather than PE. Statistically non-significant differences in secondary and safety endpoints were observed, with two major bleeding events in each arm. In conclusion, no DVT developed in COVID-19 patients hospitalized in general wards, independently of enoxaparin dosing used for thromboprophylaxis. Pulmonary artery occlusions developed only in the o.d. group. Our trial is underpowered and with few events.

**REGISTRATION:** ClinicalTrials.gov Identifier: NCT04366960

**Ethics Commettee approvation number:** 75/2020

## INTRODUCTION

The American Society of Haematology 2018 guidelines^1^ recommend that hospitalized medical patients with severe respiratory disease should receive prophylaxis with low dose unfractioned heparin (UH), low molecular weight heparin (LMWH), or fondaparinux to reduce their risk of venous thromboembolism (VTE). COVID-19 is a disease associated with SARS-CoV-2 infection, which in hospitalized patients should be treated with prophylactic anticoagulation, because they are immobilized and commonly develop interstitial pneumonia and acute respiratory distress syndrome (ARDS),^2^ which expose them to high VTE risk.^3^ However, mostly based on early observational studies, the general perception is that the VTE risk is particularly high in COVID-19; as a consequence, hospitalized COVID-19 patients are often treated with higher doses of LMWH or UFH than recommended.^4-6^ This practice does not take into due account the obvious risk of major, potentially fatal bleeding that is associated with high doses, in the absence of any evidence of higher efficacy compared to standard doses.^7,8^ Moreover, whether or not VTE risk is particularly high in COVID-19 is uncertain and difficult to assess in the absence of well-controlled studies. In a recently published meta-analysis, including studies that systematically screened patients for deep vein thrombosis (DVT) who were being treated with low-dose UH or LMWH for VTE prophylaxis, the pooled prevalence of DVT among 258 COVID patients in medical wards was 4·57% (0·00-19·84), comparable to that observed in non-COVID-19 medical patients enrolled in randomized clinical trial (RCT) of thromboprophylaxis [3·64% (1·96-5·79; p=0·789)]. Conversely, the pooled prevalence of pulmonary embolism (PE) in COVID-19 was 2.55% (0·00-9·43), compared to only 0·11% (0·00-0·31) in non-COVID-19 patients, albeit the difference did not reach statistical significance (P=0.07), likely due to insufficient statistical power.^9^ These results were considered compatible with our hypothesis that the frequent pulmonary arteries occlusions in COVID-19 are often caused by in situ thrombi, rather than PE,^10^ which was subsequently confirmed by several studies.^11-13^ It is unknown whether or not, and to what extent, high-dose UFH or LMWH can prevent the formation of pulmonary artery occlusions in COVID-19. Guidelines from several organizations still recommend standard thromboprophylaxis with low-dose LMWH or fondaparinux in acutely ill COVID-19 patients hospitalized in general wards, lacking any scientific evidence of advantages offered by higher doses.^14-17^ Given this scenario, we designed a randomized trial comparing standard prophylactic dose of subcutaneous enoxaparin (40 mg o.d.) with higher dose (40 mg b.i.d) to test whether the b.i.d. dose is more effective than the standard o.d. dose in preventing VTE in COVID-19 patients hospitalized in medical wards.

## METHODS

### Study Design

X-COVID 19 was an open-label, multicentre, prospective, controlled, randomized trial in patients admitted to medical wards with PCR-confirmed SARS-CoV-2 infection. The study was done at 9 centres in Italy. The ASST Grande Ospedale Metropolitano Niguarda and the ASST Santi Paolo and Carlo together with the operational staff of the Adivice Pharma Clinical Research Institute and the Center of Bioinformatics, Biostatistics and Bioimaging of School of Medicine and Surgery of the University of Milano Bicocca were responsible for data management, regulatory affairs and statistical analysis. The two Russian centres that initially agreed to collaborate and were included in a protocol amendment were unable to contribute and never enrolled patients.

The study protocol was approved by AIFA, the Italian Medicines Agency, and by the ethics committee of Istituto Nazionale Malattie Infettive Lazzaro Spallanzani (Rome, Italy) and accepted by other participating sites. All patients provided written informed consent. All authors had access to clinical trial data. The first and last versions of the protocol along with a summary of changes are included in the appendix. Considering the open-label design of the study no data safety and monitoring board was involved in the data reviewing for safety on an ongoing basis during the trial (considering that the formal interim analysis originally planned was not achieved). However, an independent clinical researcher and pharmacologist was responsible for data reporting to the regulatory authority.

### Participants

All patients aged >18 years admitted to hospital were eligible for inclusion. Patients were excluded when directly admitted to ICU, had an estimated creatinine clearance <15 ml/min/1·73m^2^ (CKD-EPI formula), were on anticoagulant treatment for prior indications, were on treatment with heparin at higher doses than recommended for thromboprophylaxis, were bleeding or at high bleeding risk (according to the judgement of the most responsible physician), were involved in competitive RCT exploring antithrombotic treatments or had any other condition that could either expose them at risk because of participation in the RCT or negatively affect their ability to participate in the RCT.

### Randomisation

Patients with COVID-19 (any hospitalized patient admitted to medical wards) with PCR-confirmed SARS-CoV-2 infection were randomly assigned in a 1:1 ratio to either subcutaneous 40 mg enoxaparin o.d. or 40 mg enoxaparin b.i.d. Randomization was done using an electronic web-based system using a permuted block randomization scheme with random sizes of 2, 4, 6, or 8. Enoxaparin treatment was continued until discharge.

In the first version of the protocol patients’ randomization was planned to be done within 12 hours after hospitalization; then we emended this point because the first 12 hours was considered too short a time window by the physicians, and recommended to randomize patients as early as possible. Patients were eventually randomized within a median of 6 days after admission (IQR 4-8) in the 40 mg enoxaparin o.d. and 7 days (5-10) in the 40 mg enoxaparin b.i.d.

### Procedures

The study conduct did not change the usual clinical practice. Laboratory monitoring every 3 days of the following parameters was planned: D-dimer, fibrinogen, complete blood count, LDH, PT, APTT, C-reactive protein, procalcitonin (in patients with bacterial superinfection), ferritin, CK, CK-MB, albumin, serum creatinine, AST, ALT, high-sensitivity troponin, serum bilirubin and IL-6. Patients were monitored every 7 days with compression ultrasonography (CUS) of the lower limbs for proximal DVT screening. In patients displaying signs and/or symptoms of DVT, CUS had to be performed in the same day. Proximal DVT (iliac, femoral or popliteal vein) was defined by cross-sectional vein incompressibility. Multidetector pulmonary computed tomography angiography (CTA) was performed, based on the suspicion by the treating physician of PE.

### Study outcomes

The primary endpoint was the incidence of VTE [a composite of asymptomatic or symptomatic proximal DVT diagnosed by serial CUS, and symptomatic PE diagnosed by CTA]. The secondary endpoints included: i) major adverse events (composite of overall death, VTE, use of mechanical ventilation, stroke, acute myocardial infarction and admission to ICU); ii) each single component of the primary endpoint; iii) maximum sequential organ failure assessment (SOFA); iv) levels of C-reactive protein, D-dimer, IL-6 and hs-troponin; v) Acute Respiratory Distress Syndrome (ARDS); vi) length of hospital stay; vi) changes in right ventricular function at trans-thoracic echocardiography between admission and follow-up; vii) composite of death, stroke and myocardial infarction at 30 days. Safety endpoints included: i) major bleeding events according to the International Society on Thrombosis and Haemostasis (ISTH) bleeding scale;^18^ ii) type 3 and 5 bleeding events according to the Bleeding Academic Research Consortium (BARC)^19^; iii) heparin-induced thrombocytopenia (HIT). At 30-day follow-up, patients’ dyspnea was measured according to the Borg scale.^20^ Study outcomes and their definitions are listed in **Supplement 1**. Primary and secondary outcomes were adjudicated by a clinical events committee blinded to treatment assignment.

### Sample size

The proportion of patients with VTE was expected to be 0·10 in the enoxaparin 40 mg o.d. group. With a type Iα error set to 0·05 (two-tails) and a power 1-β set to 0·8, 2712 patients were estimated to be enrolled in the trial to show a 0·03 difference.

The trial was designed with one interim analysis. This means that a preliminary test to potentially stop the trial for efficacy should have been performed after 492 out of 2712 planned subjects (246 randomized to enoxaparin 40 mg od and 246 randomized to enoxaparin 40 mg bid) had completed the trial. For the interim analysis, the proportion for the primary endpoint was estimated to be 0.07 in b.i.d. group and 0.17 in the o.d. group. With a type I error α set to 0.05 and a power 1-β set to 0.8, a total of 492 subjects would have been required.

The duration of the enrolment was projected to be 10 months, with the total duration of the study set at 13 months for follow-up, data collection, checking and analysis. However, at the end of 10 months, only 189 patients had been enrolled. Considering the enrollment constraints and the very low likelihood of reaching the expected sample size, the steering committee, blinded to the results of the trial, decided to end the study on April 25, 2021. The decision was communicated to all involved investigators, local ethics committees and AIFA.

### Statistical Analysis

Data were analysed according to the intention-to-treat principle. A test for difference of proportions was carried out to compare in-hospital composite events among patients randomized to the 2 groups. Absolute Risk Reduction (ARR) and risk ratio (RR) were estimated, as appropriate, to quantify the risk of enoxaparin o.d. compared to enoxaparin b.i.d., with 95% confidence intervals (CI). Confidence intervals were calculated with asymptotic and exact method based on the binomial distribution.

Maximum SOFA score and laboratory examinations were compared by the t-test or Wilcoxon rank-sum test, as appropriate. For the secondary outcomes there was no allowance for multiplicity. All tests were two-sided; a p-value <0·05 was considered statistically significant. The STATA version 14 (Stata Corp., College Station, TX) was used for analyses.

## RESULTS

Between April 30, 2020 and April, 25 2021, 3,550 provisionally-eligible patients were admitted in the general wards of the participating centres. Only 186 patients were randomized, mostly because of the burden of the pandemic on daily clinical practice, the treating physicians’ personal beliefs on the most appropriate anticoagulation regimen for each single patient and challenges in performing CUS systematically. Two patients were excluded because they were included in other studies requiring exclusive enrolment and 1 because positivity for SARS-Cov-2 infection was not confirmed by a second test. Therefore, 183 patients were finally included in the primary analysis (**Figure 1**). The two study groups were well-balanced for baseline clinical and laboratory characteristics (**Tables 1-3**). The median duration of treatment was comparable: 7 (IQR 4-13) days in the 40 mg o.d. group, 9 (IQR 6-13) days in the 40 mg b.i.d group, p=0·17. Treatment was changed by the treating physicians due to intercurrent clinical events in 7 (40 mg o.d.) and 4 (40 mg b.i.d) patients: in the o.d. group, enoxaparin was increased in 5 patients and switched to fondaparinux or a vitamin K antagonist in 2; in the b.i.d group, enoxaparin was discontinued in 1, increased in 2 and decreased to o.d. in 1 (**Figure 1**). Thus, since these dose adjustments were the consequence of the patient’s outcome, they could not be considered as protocol violations. For this reason, we did not compute and add the per-protocol analysis.

**Table 1.** Baseline characteristics of the primary intention to treat analysis population in a study of the effects of enoxaparin 40 mg o.d. vs 40 mg b.i.d. in patients with COVID-19 hospitalized in general wards.

| Variables | All patients (n=183) | Enoxaparin 40 mg o.d. group (n=92) | Enoxaparin 40 mg b.i.d. group (n=91) |
| --- | --- | --- | --- |
| **Medical History** |  |  |  |
| Age, years | 59 (51-72) | 59 (48-72) | 60 (53-73) |
| Male sex | 115 (62.8%) | 59 (64.1%) | 56 (61.5%) |
| BMI | 25 (23-28) | 25 (23-28) | 26 (24-28) |
| History of smoke | 29 (15.8%) | 13 (14.5%) | 16 (17.5%) |
| COPD | 10 (5.5%) | 3 (3.3%) | 7 (7.7%) |
| Hypertension | 66 (36.1%) | 29 (31.5%) | 37 (40.7%) |
| Diabetes | 25 (13.7%) | 12 (13.0%) | 13 (14.3%) |
| Dyslipidemia | 15 (8.5%) | 8 (9.0%) | 7 (8.0%) |
| Prior Stroke | 5 (2.7%) | 3 (3.3%) | 2 (2.2%) |
| Peripheral vascular disease | 7 (3.8%) | 3 (3.3%) | 4 (4.4%) |
| Chronic kidney disease | 3 (1.6%) | 3 (3.3%) | 0 |
| History of cancer | 13 (7.1%) | 8 (8.7%) | 5 (5.4%) |
| History of immunological disease | 13 (7.1%) | 7 (7.6%) | 6 (6.7%) |
| History of neurological disease | 8 (4.4%) | 4 (4.4%) | 4 (4.3%) |
| History of heart failure | 3 (1.6%) | 1 (1.1%) | 2 (2.2%) |
| Prior PCI | 5 (2.7%) | 3 (3.3%) | 2 (2.2%) |
| Prior CABG | 3 (1.6%) | 1 (1.1%) | 2 (2.2%) |
| Antiplatelet therapy | 26 (14.2%) | 12 (13.0%) | 14 (15.4%) |
| ACE-inhibitors | 28 (16.7%) | 11 (13.4%) | 17 (19.8%) |
| **Clinical Presentation** |  |  |  |
| Fever | 145 (79.2%) | 76 (82.6%) | 69 (75.8%) |
| Myalgia | 52 (28.4%) | 32 (34.8%) | 20 (22.0%) |
| Asthenia | 79 (43.2%) | 40 (43.5%) | 39 (42.9%) |
| Headache | 23 (12.6%) | 13 (14.1%) | 10 (11.0%) |
| Olfactory and Gustatory dysfunction | 27 (14.7%) | 14 (15.2%) | 13 (14.3%) |
| Cough | 102 (55.7%) | 51 (55.4%) | 51 (56.0%) |
| Dyspnea | 101 (55.2%) | 55 (59.8%) | 46 (50.5%) |
| Gastrointestinal symptoms | 50 (27.3%) | 24 (26.1%) | 26 (28.6%) |
| MBP | 92 (85-100) | 90 (85-97) | 94 (86-100) |
| Heart rate | 80 (70-90) | 78 (70-90) | 80 (72-90) |
| Respiratory rate | 18 (18-22) | 20 (18-22) | 18 (17-22) |
| pO<sub>2</sub><sup>a</sup> | 75 (65-94) | 75 (66-96) | 76 (65-89) |
| SatO<sub>2</sub><sup>a</sup> | 97 (94-98) | 96 (94-98) | 97 (95-98) |

Values are expressed as numbers (%) and medians (interquartile ranges). Differences were not statistically significant between the 2 study groups. Abbreviations: o.d., once daily; b.i.d., bis in die; BMI, Body Mass Index; COPD, Chronic Obstructive Pulmonary Disease; PCI, Percutaneous Coronary Intervention; CABG, Coronary Artery Bypass Graft; ACE, Angiotensin Converting Enzyme; MBP, Mean Blood Pressure. <sup>a</sup>With a median (IQR) FiO<sub>2</sub> of 28 (21-40).

**Table 2.** Baseline laboratory variables of the primary intention to treat analysis population in a study of the effects of enoxaparin 40 mg o.d. vs 40 mg b.i.d. in patients with COVID-19 hospitalized in general wards.

| **Variables** | **Reference ranges** | **All patients (n=183)** | **Enoxaparin 40 mg o.d. group (n=92)** | **Enoxaparin 40 mg b.i.d. group (n=91)** |
| --- | --- | --- | --- | --- |
| D-dimer, µg/mL | 0-0.57 | 0.35 (0.19-0.71) | 0.32 (0.18-0.71) | 0.36 (0.19-0.68) |
| IL-6, pg/ml | 0-16.4 | 15 (5-29) | 13 (5-26) | 21 (4-32) |
| CRP, mg/dL | 0-5 | 5.7 (2.3-12.3) | 6.0 (2.3-12.3) | 5.6 (2.3-12.4) |
| Procalcitonin, ng/ml | 0-0.5 | 0.07 (0.03-0.19) | 0.07 (0.03-0.41) | 0.07 (0.04-0.19) |
| Hb, g/dL | 12-16 | 13.6 (12.5-14.5) | 13.3 (12.1-14.3) | 13.7 (12.9-14.6) |
| Leukocytes, 10<sup>9</sup>/L | 4-10 | 6.2 (4.7-8.4) | 5.9 (4.1-7.6) | 6.9 (5.4-9.0) |
| Lymphocytes, 10<sup>9</sup>/L | 1-9 | 1.01 (0.73-1.04) | 1.03 (0.68-1.04) | 0.98 (0.81-1.43) |
| Platelet count, 10<sup>9</sup>/L | 140-440 | 202 (161-153) | 195 (163-235) | 211 (161-284) |
| INR | 0.9-1.2 | 1.08 (1.02-1.16) | 1.06 (1.02-1.15) | 1.09 (1.01-1.18) |
| aPTT, s | 24-38 | 29.9 (27.6-32.7) | 30.7 (28.4-32.9) | 28.4 (26.1-31.6) |
| Creatinine, mg/dL | 0.51-1.17 | 0.89 (0.7-1.1) | 0.88 (0.7-1.1) | 0.9 (0.7-1.1) |
| Urea, mg/dL | 18-48 | 32 (23-43) | 28 (22-40) | 35 (24-45) |
| AST, U/L | 0-40 | 35 (24-45) | 36 (26-46) | 34 (24-40) |
| ALT, U/L | 3-45 | 29 (20-46) | 29 (21-52) | 29 (19-36) |
| Bilirubin, mg/dL | 0.25-1 | 0.46 (0.34-0.63) | 0.43 (0.31-0.61) | 0.52 (0.38-0.63) |
| HS-Troponin T, ng/L | 0-14 | 7 (4-14) | 5 (3-9) | 7 (5-15) |
| NT-proBNP, ng/L | 0-155 | 107 (90-225) | 152 (81-270) | 99 (45-108) |
| BNP, pg/ml | 0-100 | 37 (25-105) | 35 (25-105) | 47 (28-122) |
| Albumin, g/dL | 3.6-4.8 | 3.5 (3.3-4) | 3.5 (3.2-3.9) | 3.6 (3.3-4) |
| Ferritin, ng/ml | 30-400 | 445 (245-931) | 505 (178-1099) | 414 (248-778) |
| Fibrinogen, mg/dL | 180-350 | 557 (503-665) | 590 (488-672) | 575 (522-652) |

Data are expressed as medians (interquartile ranges). Differences were not statistically significant between the 2 study groups, except for aPTT values (p=0.003)
Abbreviations: o.d., once daily; b.i.d., bis in die; IL-6, Interleukin 6; CRP, C Reactive Protein; Hb, Haemoglobin; INR, International Normalized Ratio; aPTT, activated Partial Thromboplastin Time; AST, Aspartate Aminotransferase; ALT, Alanine Aminotransferase; HS-Troponin, High-Sensitivity Troponin; NT-proBNP, N-terminal prohormone of Brain Natriuretic Peptide; BNP, Brain Natriuretic Peptide.

**Table 3.** Concomitant pharmacological treatments and respiratory support of the primary intention to treat analysis population in a study of the effects of enoxaparin 40 mg o.d. vs 40 mg b.i.d. in patients with COVID-19 hospitalized in general wards.

| **Variables** | **Enoxaparin 40 mg o.d. (n=92)**<br>(%, 95% confidence interval) | **Enoxaparin 40 mg b.i.d. (n=91)**<br>(%, 95% confidence interval) |
| --- | --- | --- |
| ***Concomitant pharmacological treatment*** |  |  |
| Antibiotic | 20 (21.7%) (95% CI 14%-31%) | 16 (17.6%) (95% CI 10%-27%) |
| Antiviral<sup>a</sup> | 23 (25.0%) (95% CI 16%-35%) | 19 (20.9%) (95% CI 13%-30%) |
| Remdesivir | 21 (22.8%) (95% CI 15%-32%) | 19 (20.9%) (95% CI 13%-30%) |
| Corticosteroids | 41 (44.6%) (95% CI 34%-55%) | 43 (47.2%) (95% CI 36%-58%) |
| Tocilizumab | 1 (1.1%) (95% CI 0.02%-5.9%) | 1 (1.1%) (95% CI 0.02%-5.9%) |
| Baricitinib | 0 | 1 (1.1%) (95% CI 0.02%-5.9%) |
| ***Concomitant Respiratory Support*** |  |  |
| Nasal cannula | 52 (56.5%) (95% CI 45%-67%) | 51 (56.0%) (95% CI 45%-66%) |
| High-flow nasal cannula | 12 (13.0%) (95% CI 7%-22%) | 17 (18.7%) (95% CI 11%-28%) |
| NIV | 11 (11.9%) (95% CI 6%-20%) | 17 (18.7%) (95% CI 11%-28%) |

Abbreviations: o.d., once daily; b.i.d., bis in die; NIV, Non-Invasive Ventilation.
<sup>a</sup>Sofosbuvir in 1 patient 40 mg od, ganciclovir in 1 patient 40 mg od

**Figure 1.**
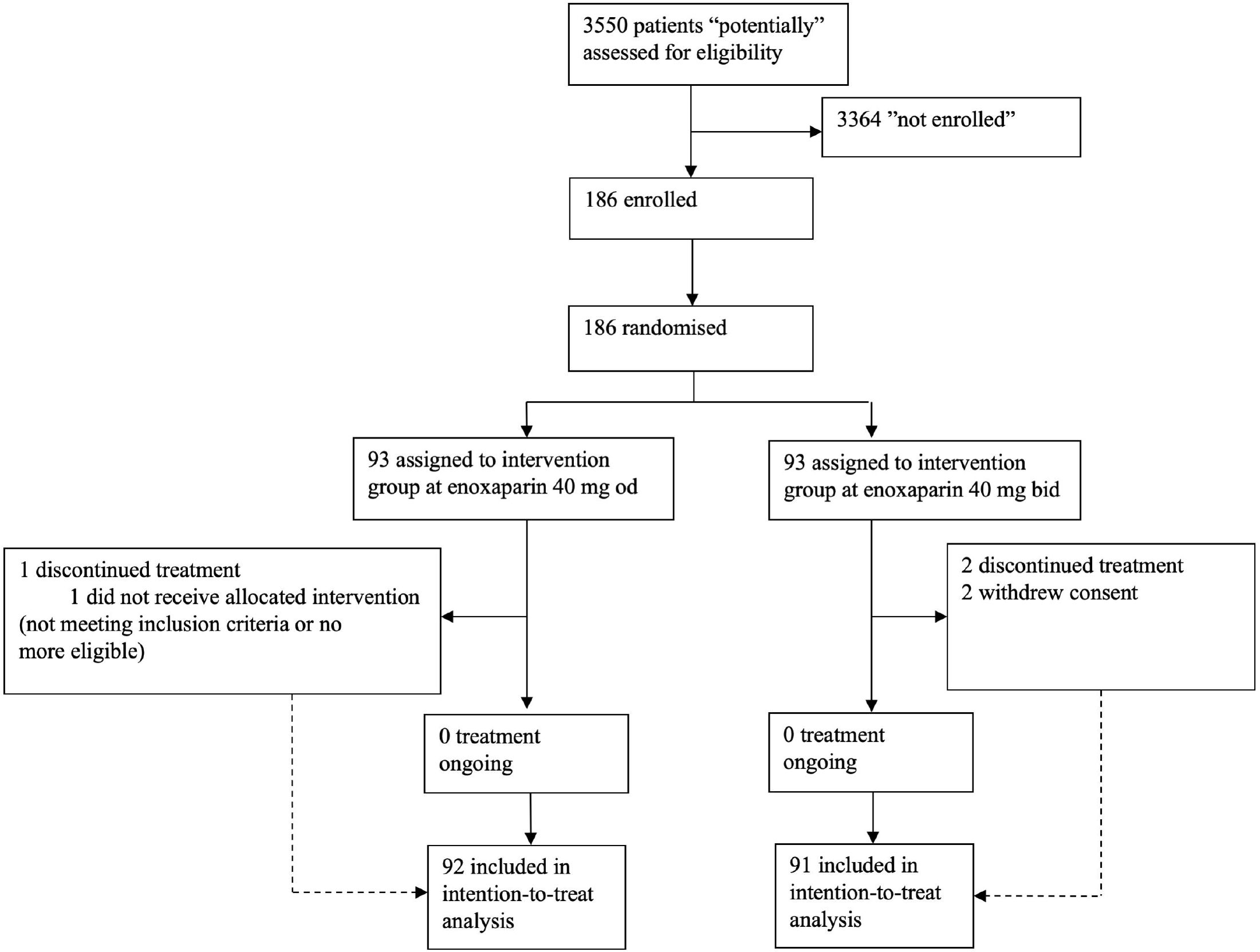
CONSORT flow diagram of the study.

### Efficacy Outcome

The primary efficacy outcome occurred in 6 patients (6·5%: 0 DVT, 6 PE) in the 40 mg o.d group and 0 patients in the 40 mg b.id group (ARR 6·5, 95% CI, 1·5-11·6) (**Table 4**). PE developed 1,3,5,6,12 and 16 days after randomization; the filling defects of pulmonary arteries were bilateral in 2, non-occlusive in 2, located in sub-segmental pulmonary arteries in 2, segmental pulmonary arteries in 3 and in a lobar artery in 1, all co-localized with inflammatory lesions of the lungs. DVT screening by CUS was assessed at baseline in 164 patients and repeated in 114. There were no statistically significant differences between patients with PE and those without, related to BMI and age (not shown). All patients with PE were discharged alive; 1 required admission to ICU for mechanical ventilation. The incidence of the secondary composite endpoint of overall death, VTE, use of mechanical ventilation, stroke, acute myocardial infarction and admission to ICU was similar: 12 patients (13%) and 9 patients (9·9%) in the enoxaparin o.d. and b.i.d. groups (**Table 4**). Death occurred in 1 patient (1·1%) in the 40 mg o.d group and 5 patients (5·5%) in the 40 mg b.id group (ARR -4·4, 95% CI, -9·5-0·7; RR 0·19, 95% CI 0·02-1·66). No statistically significant differences were observed also in the other secondary outcomes (**Table 4**).

**Table 4.** Primary, secondary, and safety in-hospital outcomes in the prespecified primary intention to treat analysis in a study of the effects of enoxaparin 40 mg o.d. vs 40 mg b.i.d. in patients with COVID-19 hospitalized n general wards.

| Outcome | Enoxaparin 40 mg<br>o.d. group<br>(n=92) | Enoxaparin 40 mg<br>b.i.d. group<br>(n=91) | Absolute risk<br>reduction |
| --- | --- | --- | --- |
| **Primary Outcome** |  |  |  |
| VTE | 6 (6.5%) | 0 | 6.5 (1.5-11.6) |
| **Secondary Outcome** |  |  |  |
| Incidence of in-hospital major complications, defined as the composite of death, VTE, use of mechanical ventilation, stroke, acute myocardial infarction and admission to an ICU | 12 (13.0%) | 9 (9.9%) | 3.1 (-6.0-12.4) |
| Pulmonary Embolism<sup>a</sup> | 6 (6.5%) | 0 | 6.5 (1.5-11.6) |
| DVT | 0 | 0 | NA |
| All-cause death | 1 (1.1%) | 5 (5.5%) | -4.4, (-9.5-0.7) |
| Stroke (ischemic) | 1 (1.1%) | 0 | 1.1 (-1.3- 3.2) |
| Composite of death, stroke and myocardial infarction at 30 days | 3 (3.5%) | 5 (5.9%) | -2.4 (-8.7-4.0) |
| Highest SOFA score | 2 (1-2) | 1 (1-2) | NA |
| SARS-CoV-2-related ARDS | 0 | 2 (3.4) | -3.4 (-8.0-1.2) |
| Mechanical ventilation | 6 (6.5%) | 5 (5.5%) | 1.0 (-5.8-7.9) |
| Length of hospital stay | 8 (6-16) | 11 (7-14) | NA |
| **Safety Outcome** |  |  |  |
| Major Bleedings (ISTH) | 1 (1.1%) | 1 (1.1%) | -0.01 (-3.0-3.0) |
| Bleedings, BARC type 3 or 5 | 1 (1.1%) | 1 (1.1%) | -0.01 (-3.0-3.0) |
| HIT | 0 | 0 | NA |

There were no cases of acute coronary syndrome. No statistically significant changes in in laboratory biomarkers were observed (not shown); too few echocardiographic evaluations have been performed to allow any meaningful analysis.
<sup>a</sup>All filling defects of the pulmonary arteries are defined as Pulmonary Embolism, although some were likely caused by local thrombi (see text for details).
Abbreviations: DVT: deep vein thrombosis; VTE, Venous ThromboEmbolism; NA, non applicable; ICU, Intensive Care Unit; SOFA, sequential organ failure assessment; ARDS: Acute Respiratory Distress Syndrome; ISTH: International Society on Thrombosis and Haemostasis classification; BARC: Bleeding Academic Research Consortium (BARC); HIT, heparin-induced thrombocytopenia.

### Safety Outcomes

There were 2 ISTH major bleeding events (type 5 and 3a BARC): 1 in the enoxaparin o.d. group (1·1%) and a fatal one in the enoxaparin b.i.d. group (1·1%) (**Table 4**); there was 1 ISTH minor bleeding (BARC 2) in the enoxaparin b.i.d. group; no cases of HIT were reported (**Table 4**).

### 30-day Follow up

At day 30, 13 patients (6·9%) were lost-to follow-up (7 in o.d. and 6 in b.i.d.). The 30-day composite of overall death, myocardial infarction and stroke occurred in 3 patients (3·5%) in the o.d. group and 5 (5·9%) in the b.i.d group (**Table 4**). Among the 170 discharged patients with 30-day follow-up, 23 (13·5%) reported persistent dyspnea, which was moderate to severe in 9.

## DISCUSSION

In this multicentre, open-label, randomized clinical trial of patients with COVID-19 admitted to the general ward, thromboprophylaxis with 40 mg o.d. enoxaparin was associated with higher incidence of the primary endpoint of VTE (proximal DVT and/or PE), compared to 40 mg b.i.d. enoxaparin. However, only the incidence of PE was different in the two treatment arms (6 events in the o.d. arm, 0 in the b.i.d. arm), while no proximal DVT was diagnosed in any of the 183 enrolled patients, independently of their treatment allocation. One patient in the enoxaparin b.i.d. arm displayed distal DVT, which was not a primary endpoint of our study. Although the small sample size of the study does not allow firm conclusions on the different efficacy of the 2 enoxaparin dose regimens, it seems reasonable to consider that both tested enoxaparin regimens (40 mg o.d. and 40 mg b.i.d.) are effective in lowering the DVT risk in COVID-19 patients hospitalized in general wards, as none of the patients displayed DVT.

These data are compatible with the results of a recent, large observational study^21^ and the demonstration by a meta-analysis of observational studies that the incidence of DVT in COVID-19 patients hospitalized in general wards and undergoing pharmacologic thromboprophylaxis is not extremely high, contrary to what had been initially surmised, but comparable to that of other medical patients at risk.^9^ However, our data on the incidence of PE are apparently in contrast with those on DVT prevention. Indeed, we observed PE in 6 patients, all treated with enoxaparin 40 mg o.d., despite the absence of DVT. This observation is difficult to reconcile with the common definition of VTE, which considers PE as a potential complication of DVT that occurs when thrombi in a deep vein break loose and travel through the bloodstream to the lungs. Although it does occasionally occur to diagnose PE in the absence of detectable DVT, this happens in only a minority of patients.^13,22^

A dramatic discrepancy between incidences of PE and DVT has already been observed in COVID-19 patients, which led to hypothesize that in many instances pulmonary artery occlusions in these patients should actually be interpreted as manifestation of local pulmonary thrombi, driven by pulmonary inflammation.^13^ Indeed, several reports later demonstrated the presence of thrombi in the pulmonary vasculature,^10-12^ generated through a thrombo-inflammatory mechanism.^23,24^ Further confirmation came from a multicentre observational study^21^ and a meta-analysis showing that the ratio between the incidences of PE and DVT was higher among COVID-19 patients than in non-COVID-19 medical patients at risk.^9^ Based on this background we tend to believe that perhaps the majority of CTA images of pulmonary artery filling defects in our 6 patients were caused by local thrombi rather than by PE. This hypothesis is corroborated by the observation that in 5 patients the filling defects affected segmental or subsegmental arteries,^24^ in 2 patients were not completely occlusive and, in all patients, colocalized with inflammatory lesions of the pulmonary parenchyma.

Whether the beneficial effect of double-dose enoxaparin is mediated by a more sustained anticoagulation, or by one or more of the pleiotropic effects of heparin (including anti-inflammatory, immunomodulatory, anti-viral and anti-complement) cannot be ascertained based on the results of our study. However, given the small number of enrolled patients, the possibility of a chance effect cannot be formally ruled out, despite the statistical significance of the observed differences.

Whatever the mechanism of action, enoxaparin b.i.d. was not efficient enough to improve the general outcome of our patients, although also this finding could have been negatively affected by lack of statistical power, due to the small sample size. The incidence of safety outcomes, in particular of bleeding complications, was also not different among the two study groups.

The plasma D-dimer levels, which are considered a risk marker for VTE and poor outcome in COVID-19 patients,^2^ were not very high in our patients and did not associate with the incidence of the endpoints of the study. Previous RCTs compared the effects of different doses of anticoagulation in COVID-19 patients, who, like those in our study, had been enrolled independently of their D-dimer levels. The INSPIRATION open-label trial, which randomized 600 COVID-19 patients hospitalized in ICU to intermediate-dose (1 mg/kg o.d.) versus standard-dose (40 mg o.d.) enoxaparin, did not show any reduction in a composite of venous or arterial thrombosis, treatment with extracorporeal oxygenation, or mortality at a follow-up of 30^11^ and 90 days.^25^ Adjudicated VTE (without distinction between DVT and PE) was comparable between the two groups (about 3%). There were 7 (2·5%) major bleeding events in the intermediate-dose group and 4 (1·4%) in the standard-dose prophylactic anticoagulation (odds ratio, 1·83 [1-sided 97·5% CI, 0·00-5·93]), with severe thrombocytopenia occurring only in 6 patients assigned to the intermediate-dose group.^8^

The AntiCoagulaTIon cOroNavirus (ACTION) trial compared therapeutic anticoagulation (in-hospital oral rivaroxaban 20 mg or 15 mg daily for stable patients, or initial subcutaneous enoxaparin 1 mg/kg b.i.d., or intravenous unfractionated heparin doses to achieve a 0·3-0·7 IU/mL anti-Xa concentration for clinically unstable patients, followed by rivaroxaban to day 30) with standard prophylactic doses of enoxaparin or UH. There were no statistically significant differences in the primary efficacy outcome (hierarchical of mortality, duration of hospitalization, and duration of oxygen use). Like in our study, the global incidence of DVT was rather low (n=10, 2%) while that of PE was relatively higher (n=20, 4%): interestingly, although the incidence of DVT was the same in each arm (n=5, 2%), that of PE tended to be higher in the prophylactic anticoagulation arm (n=13, 4%) than in the therapeutic anticoagulation arm (n=7, 2%), although the difference was not statistically significant. The incidence of major or clinically-relevant non-major bleeding was higher among patients randomized to therapeutic anticoagulation (RR 3·64; 95% CI 1·61–8·27).^26^

In the more recently published Multiplatform RCT, the reported incidence of DVT in non-critically ill patients was 0·67% (7/1046) in patients treated with standard thromboprophylaxis compared to 0·51% (6/1180) in patients in the therapeutic anticoagulation arm, whereas the incidence of PE was 1·82% versus 0·85%, respectively.^27^ Similar results were obtained in critically ill patients, in whom the incidence of DVT was 1·07% (6/559) in patients treated with standard thromboprophylaxis compared to 1·13% (6/530) in patients in the therapeutic anticoagulation arm, whereas the incidence of PE was 7·5% versus 2·4%, respectively. Therapeutic anticoagulation improved survival without organ support in non-critically ill patients,^27^ but not in critically ill patients.^28^ Survival until hospital discharge occurred in the 92·7% of the patients in the therapeutic dose anticoagulation and in 91·8% in the usual care thromboprophylaxis: adjusted difference in risk 1·3 (95% credible interval -1·1 to 3·2), adjusted odds ratio 1·21 (95% credible interval 0·87 to 1·68), with a probability of effect of therapeutic anticoagulation of 87·1%. A possible explanation for the different mortality effect compared to our trial could be the different regimen in the active arm (intermediate versus therapeutic anticoagulation) and different severity of COVID-19, with almost 30% of patients being in non-invasive respiratory support in our trial compared to the about 4% of the multiplatform trial.

Therefore, these RCTs in COVID-19 patients, similarly to the results of our study, showed that: 1) the incidence of DVT was reasonably low both in patients treated with standard prophylactic doses of anticoagulation and patients treated with higher doses; 2) the incidence of pulmonary artery occlusions was higher than that of DVT and tended to be higher in patients treated with prophylactic doses, compared to patients treated with higher doses of anticoagulants. In general, high doses anticoagulants did not improve the general clinical outcomes of the patients, with the only exception on non-critically ill patients enrolled in the Multiplatform RCT.

Our trial has some limitations. It is underpowered for primary endpoint compared to the originally planned sample size. Randomisation during the COVID-19 pandemic has been really challenging in our country. Randomisation implies a clinical equipoise of the clinician participating in the study, which is defined as “the point where we are equally poised in our beliefs between the benefits and disadvantages of a certain treatment modality. … At this point we are agnostic or resting on the fulcrum of a preference”.^29^ Among 3550 provisionally eligible patients, clinicians excluded 3364, in many cases due to the lack of personal equipoise. Clinical and individual equipoise should always be integrated in the eligibility process to understand the generalizability of a trial; other examples have been reported in literature underlying how personal believes can significantly affect the conduct of a trial.^30^Another limitation is that a minority of enrolled patients failed to undergo all the planned systematic CUS evaluations, thus some asymptomatic DVT events could have been missed.

None of the non-critically ill patients with SARS-COV2 infection enrolled in our study displayed DVT within 30-day follow-up, independently of their assignment to treatment with 40 mg o.d. or 40 mg b.i.d. We think that, despite the small number of patients enrolled, our finding (together with those of recent RCTs) represents evidence that the incidence of DVT is reasonably low in these patients when they are treated prophylactically with enoxaparin and that higher doses are not necessary. The observation that intermediates dose enoxaparin (40 mg b.i.d.) protects from the occurrence of pulmonary artery occlusions better than the standard 40 mg o.d. dose is certainly a less solid observation, in consideration of the low number of enrolled patients in each arm. However similar findings in COVID-19 patients hospitalized in ICU reported in other trials support our observation. The generalizability of our results is hampered by the small sample size of the trial. Planned metanalytical studies, which will also include our study, will hopefully overcome this limitation.

## Data Availability

All data produced in the present study are available upon reasonable request to the authors

## Author Contributions

Nuccia Morici had full access to all of the data. She takes responsibility for the integrity of the data and the accuracy of the data analysis.

### Concept and design

Marco Cattaneo and Nuccia Morici.

### Acquisition of data

GianMarco Podda, Simone Birocchi, Luca Bonacchini, Marco Merli, Michele Trezzi, Gianluca Massaini, Marco Agostinis, Giulia Carioti, Francesco Saverio Serino, Gianluca Gazzaniga, Daniela Barberis.

### Analysis of data

Marco Cattaneo, Nuccia Morici, Laura Antolini, Maria Grazia Valsecchi, Daniela Barberis.

### Interpretation of data

Marco Cattaneo, Nuccia Morici, Maria Grazia Valsecchi, Laura Antolini.

### Drafting of the manuscript

Marco Cattaneo, Nuccia Morici.

### Critical revision of the manuscript for important intellectual content

GianMarco Podda, Simone Birocchi, Luca Bonacchini, Marco Merli, Michele Trezzi, Gianluca Massaini, Marco Agostinis, Giulia Carioti, Francesco Saverio Serino, Gianluca Gazzaniga.

### Statistical analysis

Nuccia Morici, Maria Grazia Valsecchi, Laura Antolini.

### Administrative, technical, or material support

Daniela Barberis

### Other - monitoring of the study progress, supporting patient recruitment, data clarifications, and data entry

Gianluca Gazzaniga, Daniela Barberis, Giulia Carioti.

## Data Sharing

Data sharing policy has been adopted. Data will become available to interested investigators, upon submission of a reasonable research request, approved by the chief investigators of the trial.

## Acknowledgments

The study was supported only in part by research funding from Italfarmaco, which gave a contribution to allow the activation of Russian centers (I.M. Sechenov First Moscow State Medical University, Makatsariya Alexander; Central Clinical Hospital of the Presidential Department of Russian Federation, Nikita Lomakin) along with a study grant. Unfortunately, the Russiam centers had never started enrolment.

We thank all the following for supporting patients recruitment: Elena Toso, MD Francesca Casadei, MD Silvia Bondini, MD Elena Meneghini MD (ASST Grande Ospedale Metropolitano Niguarda); Sara Irene Bonelli, MD Massimo Di Pietro, MD Gregorio Basile, MD, Magdalena Viscione, MD (Malattie Infettive II, Azienda Usl Toscana CentrO, Ospedale San Jacopo Pistoia), Pierfrancesco Frosini, MD Andrea Santoro, MD (Chirurgia Vascolare, Azienda Usl Toscana Centro, Ospedale San Jacopo Pistoia), Franco Cipollini, MD Nicola Nesti, MD (Medicina Interna II, Azienda Usl Toscana Centro, Ospedale San Jacopo Pistoia).

We thank Dr Sergio Leonardi for advices in study conduct.

We thank Dr Gabriele Crimi and prof Stefano De Servi for blinded outcome adjudication.

## Authors/XCOVID-19 Investigators

GianMarco Podda, Simone Birocchi (ASST Santi Paolo e Carlo, Milan, N=64), Luca Bonacchini, Gianluca Gazzaniga, Marco Merli (ASST Grande Ospedale Metropolitano Niguarda, Milan, N=50), Michele Trezzi, Gianluca Massaini (AUSL Toscana Centro, Ospedale San Jacopo, Pistoia, N=27) Marco Agostinis, Giulia Carioti (Ospedale San Carlo Borromeo, ASST Santi Paolo e Carlo, Milano, N=26), Francesco Saverio Serino (ASL 4 Veneto, Covid Hospital, Jesolo, N=11), Paolo Righini (IRCCS Policlinico San Donato, Milan, N=6), Paolo Bonfanti (ASST Monza, Ospedale di Monza, Monza, N=3), Annamaria Municinò, Alessandro Rollero (ASL 3 Genovese, Genova, N=2), Italo Porto, Stefano Giovinazzo (IRCCS Ospedale Policlinico San Martino, Genova, N=1).

