## Appendix for "Enoxaparin for thromboprophylaxis in hospitalized COVID-19 patients: comparison of 40 mg o.d. *vs* 40 mg b.i.d. The X-COVID19 Randomized Clinical Trial"

X-COVID19 multicentre Randomized Clinical Trial

### Supplementary appendix

NUCCIA MORICI

### COMMITTEES AND INVESTIGATORS

*Steering Committee:*
Nuccia Morici, Marco Cattaneo

*Clinical Event Committee (CEC):*
Gabriele Crimi, Stefano De Servi

*Data Safety Monitoring Board:*
Data monitoring was managed by a data manager and an independent researcher (Dr. Arianna Pani), specialized in pharmacology, which checked the events every two months.

Dr. Enrico Ammirati, Marco Valgimigli and Stefano Savonitto has only partecipated to the trial as consultant in the planning phases of the study.

### FIRST PROTOCOL VERSION (VERSION 1.0)

**Enoxaparin for thromboprophylaxis in hospitalized COVID-19 patients: comparison of 40 mg o.d. versus 40 mg b.i.d. A randomized Clinical Trial**
**Internal Reference Number / Short title:** X-COVID 19
**Ethics Ref:**
**Date and Version No**: April 2020, Version 1.0

| | |
|---|---|
| **Chief Investigators:** | Marco Cattaneo, Dipartimento di Scienze della Salute, Università Degli Studi di Milano, Milan, Italy; Unità di Medicina 2, ASST Santi Paolo e Carlo, Milan, Italy; Nuccia Morici, ASST Grande Ospedale Metropolitano Niguarda, Milan, Italy. |
| **Investigators:** | Enrico Ammirati, Jacopo Oreglia, Alice Sacco, Claudia Marini, Massimo Puoti, Linda Guarnieri (ASST Grande Ospedale Metropolitano Niguarda, Milan, Italy), Gianfranco Aprigliano (Istituto Clinico Città Studi, Milan, Italy), Francesco Bedogni (IRCCS Policlinico San Donato, Milan, Italy), Gaetano Maria De Ferrari, Fabrizio D'Ascenzo (AOU Città della Salute e della Scienza, Presidio Molinette, Torino, Italy), Giovanni Forleo (Ospedale Sacco, Milan, Italy), Sergio Leonardi, Claudio Montalto (IRCCS Policlinico San Matteo, Pavia, Italy), Paolo Bonfanti (ASST Monza – Ospedale di Monza, Monza, Italy), Ilaria Battistoni, Matteo Francioni (AOU Ospedali Riuniti di Ancona, Ancona, Italy), Marco Metra, Pompilio Faggiano (ASST Spedali Civili di Brescia, Brescia, Italy), Annamaria Municinò (Ospedale Andrea Gallino, ASL 3 Genovese, Genova, Italy), Giulio Stefanini (IRCCS Istituto Clinico Humanitas, Rozzano, Milan, Italy), Stefano Savonitto (ASST Lecco, Ospedale A. Manzoni, Lecco, Italy) |
| **Sponsor:** | ASST Grande Ospedale Metropolitano Niguarda, Milan, Italy |
| **Funder:** | None |
| **Chief Investigator Signature:** | Marco Cattaneo, Nuccia Morici |
| **Statistician Signature:** | Gianfranco Alicandro |

**Confidentiality Statement:** This document contains confidential information that must not be disclosed to anyone other than the Sponsor, the Investigator Team, HRA, host organisation, and members of the Research Ethics Committee, unless authorised to do so.

#### TABLE OF CONTENTS

1. KEY TRIAL CONTACTS 6

2. SYNOPSIS 7

3. ABBREVIATIONS 9

4. BACKGROUND AND RATIONALE 10

5. OBJECTIVES AND OUTCOME MEASURES 11

6. TRIAL DESIGN 12

7. PARTICIPANT IDENTIFICATION 13

   7.1 Trial participants 13

   7.2 Inclusion Criteria 13

   7.3 Exclusion Criteria 14

8 TRIAL PROCEDURES 14

   8.1 Recruitment 15

   8.2 Informed Consent 15

   8.3 Screening and Eligibility Assessment 15

   8.4 Randomisation, blinding and code-breaking 16

   8.5 Baseline Assessments 16

   8.6 Subsequent Visits 16

   8.7 Discontinuation/Withdrawal of Participants from Trial Treatment 17

   8.8 Definition of End of Trial 18

9. COMPLIANCE WITH TRIAL TREAMTENT 18

10. ACCOUNTABILITY OF TRIAL TREATMENT 18

11. CONCOMITANT MEDICATION 18

12. POST-TRIAL TREATMENT 18

13. SAFETY REPORTING 18

   13.1. Definitions 18

   13.2. Causality 19

   13.3. Procedures for Recording Adverse Events 19

   13.4. Reporting Procedures for Serious Adverse Events 20

   13.5. Expectedness 20

   13.6. SUSAR Reporting 20

   13.7. Safety Monitoring Committee 20

14. STATISTICS 20

   14.1. Description of Statistical Methods 20

   14.2. Number of Participants 21

   14.3. Populations Analysis 21

   14.4. Criteria for the Termination of the Trial 22

   14.5. Procedure for Accounting for Missing, Unused, and Spurious Data 22

14.6. Inclusion in Analysis 22
14.7. Procedures for Reporting any Deviation(s) from the Original Statistical Plan 22
15. DATA MANAGEMENT 22
15.1. Source Data 22
15.2. Access to Data 23
15.3. Data Recording and Record Keeping 23
16. QUALITY ASSURANCE PROCEDURES 23
17. SERIOUS BREACHES 23
18. ETHICAL AND REGULATORY CONSIDERATIONS 23
18.1. Declaration of Helsinki 23
18.2. Guidelines for Good Clinical Practice 24
18.3. Approvals 24
18.4. Reporting 24
18.5. Participant Confidentiality 24
18.6. Expenses and Benefits 24
19. FINANCE AND INSURANCE 24
19.1. Funding24
19.2. Insurance 24
20. PUBLICATION POLICY 25
21. REFERENCES 26

### 1. KEY TRIAL CONTACTS

| | |
|---|---|
| **Chief Investigator** | Marco Cattaneo, Dipartimento di Scienze della Salute, Università Degli Studi di Milano, Milan, Italy; Unità di Medicina 2, ASST Santi Paolo e Carlo, Milan, Italy. Phone 0250323095 - 0281844638; fax 0250323089;<br>Nuccia Morici, Unità di Cure Intensive Cardiologiche Dipartimento Cardio-toraco-vascolare, ASST Grande Ospedale Metropolitano Niguarda, Milan, Italy. Phone 0264442565, fax 0264442818, |
| **Sponsor** | ASST Grande Ospedale Metropolitano Niguarda, Milan, Italy |
| **Clinical Trials Unit** | Unità di Medicina, ASST Santi Paolo e Carlo, Milan, Italy / Malattie Infettive, ASST Grande Ospedale Metropolitano Niguarda, Milan, Italy |
| **Statisticians** | Gianfranco Alicandro, |
| **Committees** | **Comitato Etico Scientifico INMI Lazzaro Spallanzani (Ethical Committee)**<br>**Steering Committee**: Marco Cattaneo, Nuccia Morici, Enrico Ammirati, Marco Valgimigli, Stefano Savonitto. |

### 2. SYNOPSIS

| | |
|---|---|
| Trial Title | **Enoxaparin for thromboprophylaxis in hospitalized COVID-19 patients: comparison of 40 mg o.d. versus 40 mg b.i.d. A randomized Clinical Trial** |
| Internal ref. no. (or short title) | X-Covid 19 |
| Promoter | ASST Grande Ospedale Metropolitano Niguarda |
| Principal Investigator | Marco Cattaneo / Nuccia Morici |
| EudraCT Number | 2020-001708-41 |
| Clinical Phase | Phase IIIb |
| Trial Design | Open-label Multicentre Prospective Controlled Randomized Trial in patients with SARS-CoV-2 infection. Patients will be randomized 1:1 to 40 mg subcutaneous enoxaparin o.d. versus 40 mg enoxaparin b.i.d within 12 hours after hospitalization. |
| Trial Participants<br>*Inclusion criteria* | • All-comers patients aged >=18 years and admitted to hospital with laboratory-confirmed SARS-CoV-2 infection. |
| *Exclusion criteria* | • Patients admitted directly to an intensive care unit;<br>• Estimated creatinine clearance <15 ml/min/1.73m$^2$;<br>• Patients needing anticoagulant for prior indication;<br>• Patients at high bleeding risk or experiencing clinically significant bleeding;<br>• Patients involved in clinical trial protocols restricting patients from concurrent studies;<br>• Any other significant disease or disorder which, in the opinion of the investigator, may either put the participants at risk because of participation in the trial, or may influence the result of the trial, or the participant's ability to participate in the trial. |
| Planned Sample Size | A sample of 2712 subjects is required to test the superiority hypothesis with 80% power according to a two-sided 0.05 level of significance that 40 mg enoxaparin b.i.d. will decrease the primary endpoint compared to 40 mg enoxaparin o.d. An event rate of 10% was hypothesized in the control arm and 7% in the treatment arm, corresponding to a 33% relative risk reduction. |
| Treatment duration | In-hospital (at least 30 days of therapy) |
| Follow up duration | 30 days from the enrolment |
| Planned Trial Period | 6 months |
| Primary Objective | To compare the effects of 40 mg subcutaneous enoxaparin o.d. versus 40 mg enoxaparin b.i.d on the incidence of venous thromboembolism (VTE) [a composite of incident asymptomatic and symptomatic deep vein thrombosis (DVT) diagnosed by serial compression ultrasonography (CUS), and symptomatic pulmonary embolism (PE) diagnosed by CT scan], in patients with SARS-CoV-2 infection. |

| | |
|---|---|
| Secondary Objectives | • To compare the effects of 40 mg subcutaneous enoxaparin o.d. versus 40 mg enoxaparin b.i.d on the incidence of in-hospital major complications, defined as the composite of death, VTE, use of mechanical ventilation, stroke, acute myocardial infarction and admission to an intensive care in patients with SARS-CoV-2.<br>• To compare each single component of the primary endpoint between the two groups.<br>• To compare maximum sequential organ failure assessment (SOFA) score between the two groups.<br>• To compare C-reactive protein, D-dimer, IL-6 and hs-troponin levels (as % above the upper reference limit [URL]) among the two groups.<br>• To compare the incidence of SARS-CoV-2-related Acute Respiratory Distress Syndrome (ARDS) between the two groups.<br>• To compare length of hospital stay between the two groups.<br>• To compare measures of right ventricular function at trans-thoracic echocardiography or CT between admission and follow-up, whenever available. |
| Safety Endpoints | • To compare the incidence of bleeding events according to the *ISTH bleeding scale* between the two groups.<br>• To compare the incidence of bleeding events according to *BARC classification* between the two groups. Only BARC 3 and 5 will be considered for secondary endpoint.<br>• To assess the incidence of heparin-induced thrombocytopenia in the two groups. |

#### 3. ABBREVIATIONS

| | |
|---|---|
| AE | Adverse event |
| AR | Adverse reaction |
| BARC | Bleeding Academic Research Consortium |
| CI | Chief Investigator |
| CRF | Case Report Form |
| CT | Clinical Trials |
| CTA | Clinical Trials Authorisation |
| CTRG | Clinical Trials and Research Governance |
| DMC/DMSC | Data Monitoring Committee / Data Monitoring and Safety Committee |
| DSUR | Development Safety Update Report |
| GCP | Good Clinical Practice |
| GP | General Practitioner |
| IB | Investigator's Brochure |
| ICF | Informed Consent Form |
| ICH | International Conference on Harmonisation |
| IMP | Investigational Medicinal Product |
| IRB | Independent Review Board |
| PI | Principal Investigator |
| PIL | Participant/ Patient Information Leaflet |
| REC | Research Ethics Committee |
| SAE | Serious Adverse Event |
| SAR | Serious Adverse Reaction |
| SDV | Source Data Verification |
| SMPC | Summary of Medicinal Product Characteristics |
| SOP | Standard Operating Procedure |
| SOFA | Sequential Organ Failure Assessment |
| SUSAR | Suspected Unexpected Serious Adverse Reactions |
| TMF | Trial Master File |

### 4. BACKGROUND AND RATIONALE

Critically ill medical patients are at heightened risk for venous thromboembolism (VTE) and, for this reason, they are treated with prophylactic doses of low molecular weight heparin (LMWH) (for instance, 40 mg enoxaparin). Patients with COVID-19, which is caused by SARS-CoV-2 infection, are apparently at particularly high risk of VTE, as a consequence of excessive activation of the hemostatic system which, in the most severe cases, could also be associated with the formation of microthrombi and clinically relevant disseminated intravascular coagulation.[1-4]

Based on personal communications by medical doctors working mostly in Northern Italy, there are concerns about the efficacy of thromboprophylaxis with low-molecular-weight heparin (LMWH) at the standard doses in these patients, given their apparently very high risk of thromboembolic complications. Therefore, in many Northern Italian hospitals these patients are given LMWH at much higher doses than those recommended for thromboprophylaxis: in some hospitals, full anticoagulant doses (100 U/Kg b.i.d.) are being used even in the absence of evidence of ongoing thromboembolic disease. This practice does not take into due account the obvious risk of major, potentially fatal bleeding that is associated with high dosing, in the absence of any evidence of higher efficacy of high doses compared to standard prophylactic doses.

An additional rationale for the use of high dose LMWH in many hospitals is that hyper-activation of the hemostatic system in COVID-19 patients could lead to the formation of microthrombi in pulmonary vessels[5], causing further respiratory distress, and in vessels of other vital organs, causing multi-organ failure (MOF). However, there is no demonstration that this therapeutic approach decreases the risk of microangiopathic thrombosis in these patients, as well as, for that matter, in other types of patients.

Given this scenario, we designed a randomized trial comparing standard prophylactic dose of subcutaneous enoxaparin (40 mg o.d.) with higher dose (40 mg b.i.d) with the aim of testing whether high-dose thromboprophylaxis is more effective than standard dose in preventing TEV in COVID-19 patients. This study would also allow us to test whether double-dose prophylaxis with enoxaparin would favourably affect the natural history of the disease, by decreasing the incidence of organ dysfunction associated with the formation of microthrombi in vital organs.

### 5. OBJECTIVES AND OUTCOME MEASURES

| **Objectives** | **Outcome measures** | **Timepoint(s) of evaluation of these outcome measure (if applicable)** |
|---|---|---|
| **Primary Objective**<br>• To compare the incidence of hospital-associated venous thromboembolism (HA-VTE) between the two groups (40 mg o.d. vs 40 mg b.i.d subcutaneous enoxaparin) | • ***Number (percentage) of patients who will experience*** the composite event.<br><br>• ***Relative risk*** will evaluate the association between the study outcome and treatment. | 30-day |
| **Secondary Objectives**<br>• To compare the incidence of in-hospital major complications, defined as the composite of death, HA-VTE, use of mechanical ventilation, major stroke, acute myocardial infarction and admission to an intensive care between the two groups.<br><br>• To compare each single component of the primary endpoint between the two groups.<br><br>• To compare maximum sequential organ failure assessment (SOFA) score between the two groups.<br><br>• To compare C-reactive protein, D-dimer, IL-6 and hs-troponin levels (as % above the upper reference limit [URL]) among the two groups. | • ***Number (percentage) of patients who will experience*** the event. ***Relative risk*** will evaluate the association between the study outcome and treatment.<br><br>• ***Number (percentage) of patients who will experience*** the event. ***Relative risk*** will evaluate the association between the study outcome and treatment.<br><br>• Maximum sequential organ failure assessment (SOFA) score summarized by ***median and interquartile range (IQR) or mean and standard deviation*** according to variable distribution.<br><br>• Laboratory parameters will be indicated as ***median and interquartile range (IQR) or mean and standard deviation*** according to variable distribution. | 30-day<br><br>30-day<br><br>Discharge<br><br>Discharge<br><br>30-day<br><br>Discharge<br><br>30-day |

| | | |
|---|---|---|
| • To compare the incidence of SARS-CoV-2- related Acute Respiratory Distress Syndrome (ARDS) between the two groups.<br><br>• To compare length of hospital stay<br><br>• To compare measures of right ventricular function at trans-thoracic echocardiography or CT between admission and follow-up, whenever available. | • ***Number (percentage) of patients who will experience*** the event. ***Relative risk*** will evaluate the association between the study outcome and treatment.<br><br>• ***Median and interquartile range (IQR) or mean and standard deviation*** according to variable distribution.<br><br>• Echocardiographic parameters will be summarized by ***median and interquartile range (IQR) or mean and standard deviation*** according to variable distribution. | |
| **Safety endpoints**<br>• To assess the occurrence of bleeding events, according to the two classifications selected<br><br>• To assess the incidence of heparin-induced thrombocytopenia between the two groups. | • ***Number (percentage) of patients who will experience*** the event. ***Relative risk*** will evaluate the association between the study outcome and treatment.<br><br>• The incidence rate will be described across groups | During hospital stay<br><br>During hospital stay |

#### 6. TRIAL DESIGN

Prospective controlled randomized, open-label multi-center trial.

Patients with COVID-19 (any hospitalized patients) with laboratory-confirmed SARS-CoV-2 infection will be randomized 1:1 to 40 mg enoxaparin/day subcutaneous versus 40 mg enoxaparin b.i.d. within 12 hours after hospitalization. Standard dosage of Proton Pump Inhibitor (PPI) will be prescribed to each enrolled patient to reduce gastrointestinal (GI) bleeding risk. The chronic kidney disease epidemiology (CKD-EPI) formula will be used for eGFR calculation.

The study conduct will not change the usual clinical practice in terms of treatments and diagnostic procedures.

Patients will be monitored every 3 days with D-dimer, fibrinogen, complete blood count, lactate dehydrogenase (LDH), PT, APTT, C-reactive protein, procalcitonin only in the case of superinfection by a different agent, ferritin, creatine phosphokinase (CK), CK-MB, albumin, serum creatinine, aspartate transaminase (AST), alanine transaminase (ALT), high-sensitivity troponin, serum bilirubin and interleukine-6 along with other examinations according to clinical practice.

Patients will be monitored every seven days from the enrolment with CUS of the lower limbs for proximal DVT screening. For patients who display signs and symptoms of DVT, CUS will be performed as soon as possible in the same day. Proximal DVT, namely DVT of iliac, femoral and popliteal veins down to popliteal trifurcation, is defined by cross-sectional vein incompressibility or direct thrombus imaging with vein enlargement. Distal DVT might be reported if distal segments are explored.
For patients who display signs and symptoms of pulmonary embolism, a CT scan will be performed as soon as possible in the same day according to the indication of the attending physician. Multidetector CT pulmonary angiography is the method of choice for imaging the pulmonary vasculature in patients with suspected pulmonary embolism, that is defined by contrast-filling defects.
With regard to secondary objective, the following definitions apply.
All-cause mortality includes all deaths, regardless of whether the cause of death is determined. Cardiovascular death is defined as death due to atherosclerotic coronary heart disease, cerebrovascular accident or (complication of) peripheral embolization, and includes deaths due to acute MI, stroke, sudden death, non-sudden death, unwitnessed death, and procedure-related deaths. Non-cardiovascular death includes deaths due to all other causes.
Myocardial Infarction denotes the presence of acute myocardial injury detected by abnormal cardiac biomarkers [Cardiac troponin I (cTnI) and T (cTnT) increased above the 99th percentile upper reference limit (URL)] in the setting of evidence of acute myocardial ischaemia.
Stroke is defined as any new neurological deficit lasting >24 hours. CT or MRI are required to confirm the stroke and to distinguish between ischemic and hemorrhagic stroke. Strokes will be also classified as fatal or nonfatal: nonfatal stroke will be also classified as disabling and non disabling.
The total duration of the study is 6 months for enrolment, follow-up, data collection, cleaning and analysis.

### 7. PARTICIPANT IDENTIFICATION

#### 7.1 Trial participants

Patients directly admitted to hospital for suspected SARS-CoV-2 infection will be screened for randomization.

#### 7.2 Inclusion Criteria

All the following inclusion criteria will be adopted:

All-comers patients aged >=18 years and admitted to hospital with laboratory-confirmed SARS-CoV-2 infection.

##### 7.3 Exclusion Criteria

The following patients will not be enrolled:

- Patients admitted directly to intensive care unit.
- Patients with estimated creatinine clearance <15 ml/min/1.73m$^2$.
- Patients needing anticoagulant for prior indication.
- Patients at high bleeding risk or experiencing clinically significant bleeding;
- Patients involved in clinical trial protocols restricting patients from concurrent studies
- Patients with any other significant disease or disorder which, in the opinion of the Investigator, may either put the participants at risk because of participation in the trial, or may influence the result of the trial, or the participant's ability to participate in the trial.

### 8 TRIAL PROCEDURES

Eligibility will be evaluated by the attending physician after the first visit and after confirmation of the diagnosis. The patient, properly informed about the study, will be able to decide to sign the informed consent and the privacy form. The physician will obtain a detailed anamnesis and he/she will carefully check the clinical parameters needed to define tissue perfusion (as detailed by the inclusion criteria) along with body temperature. The following laboratory data will be checked: haemochrome, renal function, serum electrolytes, liver function, coagulation, blood gas analysis. Laboratory examinations will be sequentially evaluated according to the flow chart timeline (section 8.6). If the patient develops a condition needing an anticoagulation, an increase in enoxaparin dosage to 100 UI/kg (1 mg/kg) b.i.d. might be considered if indicated and in the absence of contraindications. Serial CUS will be performed, the first on admission and then every seven days. Bleeding events will be registered and graded according to the International Society on Thrombosis and Haemostasis (ISTH) bleeding scale and Bleeding Academic Research Consortium (BARC) scale. Also BARC 2 bleedings will be recorded.

Pre-specified subgroup analyses will be performed for 1) D-dimer categories (>1 <2, 2, 3, 4, 5, 6 fold the ULN of the reference range); 2) estimated creatinine clearance (< versus >60 ml/min/1.73 m2); 3) severity of the respiratory symptoms (mild versus moderate-severe defined as respiratory rate ≥30 breaths/min; arterial oxygen saturation≤93% at rest; PaO2/FiO2 ≤300 mmHg); 4) concomitant antiplatelet therapy.

The identification of adverse reactions will be continuous during hospitalisation. The data will be recorded in an eCRF.

#### 8.1 Recruitment

Patients recruitment will be performed at the Cardiology Department, Malattie Infettive and Internal Medicine, ASST Grande Ospedale Metropolitano Niguarda, Milan, Italy; Medicina, Ospedale San Paolo, ASST Santi Paolo e Carlo, Milan, Italy; Istituto Clinico Città Studi, Milan, Italy; IRCCS Policlinico San Donato, Milan, Italy; AOU Città della Salute e della Scienza, Presidio Molinette, Torino, Italy; Ospedale Sacco, Milan, Italy; IRCCS Policlinico San Matteo, Pavia, Italy; ASST Monza – Ospedale di Monza, Monza, Italy; AOU Ospedali Riuniti di Ancona, Ancona, Italy; ASST Spedali Civili di Brescia, Brescia, Italy; Ospedale Andrea Gallino, ASL 3 Genovese, Genova, Italy; IRCCS Istituto Clinico Humanitas, Rozzano, Milan, Italy; ASST Lecco, Ospedale A. Manzoni, Lecco, Italy.

#### 8.2 Informed Consent

The participant must personally sign and date the latest approved version of the Informed Consent form before any trial specific procedures.

Written and verbal versions of the Participant Information and Informed Consent will be presented to the participants detailing no less than: the exact nature of the trial; what it will involve the participant; the implications and constraints of the protocol; the known side effects and any risks involved in taking part. It will be clearly stated that the participant is free to withdraw from the trial at any time for any reason without prejudice to future care and with no obligation to give the reason for withdrawal.

The participant will be allowed as much time as wished, taking into consideration the urgency of treatments, to consider the information and the opportunity to ask the Investigator, his/her GP or other independent parties to decide whether he/she will participate in the trial. Written Informed Consent will then be obtained by means of participant dated signature and dated signature of the person who presented and obtained the Informed Consent. The person who obtained the consent must be suitably qualified and experienced, and authorized to do so by the Chief/Principal Investigator. The original signed Informed Consent will be given to the participant and a copy will be retained at the trial site.

#### 8.3 Screening and Eligibility Assessment

As clinically indicated, the screening procedures will be performed throughout demographics, medical history, concomitant medication, physical examination, laboratory tests and radiological findings (chest CT scans are encouraged over chest X-rays).

The screening procedure will be immediately followed by recruitment at arrival in the ward (patients will not be randomized in the emergency department), if allowed on basal evaluation and patient informed consent signature.

#### 8.4 Randomisation, blinding and code-breaking

This study will be a two-parallel group randomized, open-label, multi-center trial in hospitalized COVID-19 patients. Patients will be randomized by the attending physicians using an IVRS that randomizes at a ratio of 1:1 upon determination of eligibility.

A total sample of 2712 subjects is required to test the superiority hypothesis with 80% power according to a two-sided 0.5 level of significance that 40 mg enoxaparin subcutaneously bid/day (treatment arm) will decrease the primary endpoint compared to 40 mg enoxaparin/day (prophylaxis dose, that is considered the control arm as represents the standard of care for hospitalized patients that are bedridden). An absolute reduction of 3% is foreseen in the treatment arm (enoxaparin 40 mg b.i.d); an event rate of 10% was considered in the control arm (enoxaparin 40 mg o.d.) and 7% in the treatment arm.

The duration of the enrolment is 3 months from the obtaining of the approvals. The total duration of the study is 6 months for follow-up (1 months), data collection, cleaning and analysis (2 months). Randomisation will be done at the same visit as the baseline visit. There will not be a run-in period.

#### 8.5 Baseline Assessments

##### Basal evaluation (V1)

Every patient will be informed about the objectives of our study and he/she will be asked for consent to the trial. At basal visit and following ones, the physician will perform: physical examination, hemodynamic parameters and blood gas analysis check; CUS. Echocardiogram might be performed if requested by the attending physician.
The occurrence of organ dysfunction and failure, from admission through days spent in intensive care, will be assessed by the Sequential Organ Failure Assessment (SOFA) score.

#### 8.6 Subsequent Visits

After basal assessment, the following visits will be performed according to local clinical practice a check-up at discharge will be performed. Echocardiograms might be performed during hospitalization according to the indication of the attending physician. In case mechanical ventilation is required, ventilation modes and eventual variations of parameters will be recorded.
The schedule of clinical visits and assessment procedures is shown in the following flow chart.

**Flow chart**

| Visit | V1 | V2 | V2 | V3 | V4 |
|---|---|---|---|---|---|
| **Time interval** | **Basal** | **To be checked each three days** | **To be checked each seven days** | **Discharge** | **30-day from the enrolment** |
| **Informed consent** | X | | | | |
| **Time from onset symptoms to enrolment** | X | | | | |
| **Physical examination** | X | | | | |
| **Height** | X | | | | |
| **Weight** | X | | | | |
| **Anamnesis** | X | | | | X |
| **BP (s/d/m)** | X | | | | |
| **HR** | X | X | | X | |
| **CUS** | X | | X | | |
| **Hb** | X | X | | X | |
| **Creatinine** | X | X | | X | |
| **Platelets** | X | X | | X | |
| **Leucocytes** | X | X | | X | |
| **INR** | X | X | | X | |
| **Bilirubin** | X | X | | X | |
| **AST** | X | X | | X | |
| **ALT** | X | X | | X | |
| **LDH** | X | X | | X | |
| **CRP** | X | X | | X | |
| **Troponin (HS)** | X | X | | X | |
| **IL-6** | X | X | | X | |
| **D-dimer** | X | X | | X | |
| **Fibrinogen** | X | X | | X | |
| **SOFA score** | X | X | | | |

#### 8.7 Discontinuation/Withdrawal of Participants from Trial Treatment

Each participant has the right to withdraw from the trial at any time. In addition, the Investigator may discontinue a participant from the trial at any time if the Investigator considers it necessary for any reason including:

- Ineligibility (either arising during the trial or retrospectively having been overlooked at screening)
- Significant protocol deviation
- Significant non-compliance with treatment regimen or trial requirements
- An adverse event which requires discontinuation of trial treatment or results in inability to continue to comply with trial procedures
- Disease progression which requires discontinuation of trial treatment or results in inability to continue to comply with trial procedures
- Withdrawal of Consent

Withdrawal from the trial will result in exclusion of the data for that participant from analysis

The reason for withdrawal will be recorded in the eCRF.

If the participant is withdrawn due to an adverse event, the Investigator will arrange for follow-up visits or telephone calls until the adverse event has resolved or stabilised.

##### 8.8 Definition of End of Trial

The end of trial is the date of the last visit/ telephone follow up/ home visit of the last participant.

#### 9. COMPLIANCE WITH TRIAL TREAMTENT

All patients will be treated in hospital ward, so compliance will be guaranteed by the investigators.

#### 10. ACCOUNTABILITY OF TRIAL TREATMENT

The study does will involve drugs usually administrated in clinical practice.

#### 11. CONCOMITANT MEDICATION

There are not any contraindicated medications. All used drugs, as per clinical indication, will be reported in the eCRF.

#### 12. POST-TRIAL TREATMENT

At the end of the study period, all surviving patients will be treated as per current clinical practice in the participating Centers.

#### 13. SAFETY REPORTING

##### 13.1. Definitions

| | |
|---|---|
| Adverse Event (AE) | Any untoward medical occurrence in a participant to whom a medicinal product or diagnostic-interventional procedure has been administered, including occurrences which are not necessarily caused by or related to that product or procedure. |
| Adverse Reaction (AR) | An untoward and unintended response in a participant to a medicinal product which is related to any dose administered to that participant.<br>The phrase "response to an investigational medicinal product" means that a causal relationship between a trial medication and an AE is at least a reasonable possibility, i.e. the relationship cannot be ruled out.<br>All cases judged by either the reporting medically qualified professional or the Sponsor as having a reasonable suspected causal relationship to the trial medication qualify as adverse reactions. |
| Serious Adverse Event (SAE) | A serious adverse event is any untoward medical occurrence that:<br>• Results in death |

| | |
|---|---|
| | • Is life-threatening<br>• Requires inpatient hospitalisation or prolongation of existing hospitalisation<br>• Results in persistent or significant disability/incapacity<br>• Consists of a congenital anomaly or birth defect.<br><br>Other 'important medical events' may also be considered serious if they jeopardise the participant or require an intervention to prevent one of the above consequences.<br><br>NOTE: The term "life-threatening" in the definition of "serious" refers to an event in which the participant was at risk of death at the time of the event; it does not refer to an event which hypothetically might have caused death if it were more severe. |

NB: to avoid confusion or misunderstanding of the difference between the terms "serious" and "severe", the following note of clarification is provided: "Severe" is often used to describe intensity of a specific event, which may be of relatively minor medical significance. "Seriousness" is the regulatory definition supplied above.

#### 13.2. Causality

The relationship of each adverse event to trial treatment must be determined by a medically qualified individual according to the following definitions:

**Related**: The adverse event follows a reasonable temporal sequence from trial treatment start. It cannot reasonably be attributed to any other cause.

**Not Related**: The adverse event is probably produced by the participant's clinical state or by other modes of therapy administered to the participant.

#### 13.3. Procedures for Recording Adverse Events

All AEs occurring during the trial / or until the following two months that are observed by the Investigator or reported by the participant, will be recorded on the eCRF, whether or not attributed to trial treatment.

The following information will be recorded: description, date of onset and end date, severity, assessment of relatedness to trial treatment, other suspect drug or device and action taken. Follow-up information should be provided as necessary.

The severity of events will be assessed on the following scale: 1 = mild, 2 = moderate, 3 = severe.

AEs considered related to the trial treatment as judged by a medically qualified investigator will be followed either until resolution, or the event is considered stable.

It will be left to the Investigator's clinical judgment to decide whether an AE is of enough severity to require the participant's removal from treatment. A participant may also voluntarily withdraw from treatment due to what he or she perceives as an intolerable AE. If either of these occurs, the participant must undergo an end of trial assessment and be given appropriate care under medical supervision until symptoms cease, or the condition becomes stable.

##### 13.4. Reporting Procedures for Serious Adverse Events

All SAEs (other than those defined in the protocol as not requiring reporting) must be reported on the SAE reporting form to Eudravigilance within 24 hours of the Site Study Team becoming aware of the event.

Review of SAEs must be timely, taking into account the reporting time for a potential SUSAR.

##### 13.5. Expectedness

Expectedness will be determined according to the Investigators' Brochure/Summary of Product Characteristics.

##### 13.6. SUSAR Reporting

All SUSARs will be reported by the CI to the relevant Competent Authority and to the REC and other parties as applicable. For fatal and life-threatening SUSARS, this will be done no later than 7 calendar days after the investigator team is first aware of the reaction. Any additional information will be reported within 15 calendar days. Treatment codes will be un-blinded for specific participants.

##### 13.7. Safety Monitoring Committee

The aims of this committee include:

- To pick up any trends, such as increases in un/expected events, and take appropriate action
- To seek additional advice or information from investigators where required
- To evaluate the risk of the trial continuing and take appropriate action where necessary

#### 14. STATISTICS

##### 14.1. Description of Statistical Methods

All protocol-defined endpoints will be measured from the time of randomization. Descriptive statistics of baseline characteristics of patients randomized to the two groups will be reported. Continuous variables will be summarized by means, standard deviations, medians, inter-quartile ranges and minimum and maximum values. Categorical variables will be summarized by frequencies and percentages.

Data will be analysed according to the intention-to-treat principle. In addition, a per-protocol analysis of the primary endpoint, considering patients without major protocol violations (i.e. patients who crossed over), will be performed to evaluate the robustness of the data.
A test for difference of proportions will be carried out to compare in-hospital composite events among patients randomized to the two groups. A relative risk will be calculated to quantify the magnitude of the half anticoagulant effect.
For what regards continuous secondary endpoints, maximum SOFA score will be compared among the study groups through the t-test or Wilcoxon rank-sum test, according to the distribution of study endpoints. The same approach will be applied for laboratory examinations and times included in the secondary endpoints.
All tests will be two-sided and a p-value of 0.05 or less will be considered statistically significant. No adjustments for multiple testing will be applied in the analysis of secondary endpoints and in the subgroup analysis, therefore, inferences drawn from the reported p-values and 95% CIs will be considered exploratory. If a p-value is less than 0.001 it will be reported as "<0.001".

#### 14.2. Number of Participants

**Summary Statements**
The proportion for the primary endpoint is estimated to be 0.07 in the half-dose anticoagulant dose and 0.10 in the standard group. With a type I error $\alpha$ set to 0.05 and a power 1- $\beta$ set to 0.8, a total of 2712 subjects will be enrolled in the trial.

#### 14.3. Populations Analysis

For this trial, the following populations will be defined and used in the analysis and/or presentation of the data.

**Safety population:** The safety population will be classified according to the actual treatment on enoxaparin 40 mg b.i.d. or control, received.

**Intent-to-treat (ITT) population***:* Treatment classification for ITT analysis will be based on the randomized treatment.

**Per-Protocol (PP) population:** The PP population will be defined as all ITT patients who received assigned to enoxaparin 40 mg b.i.d. and will be without major protocol violations. Specifically, subjects may be excluded from PP population if any of the following criteria are met:

- Inclusion/Exclusion criteria violations;
- 

The PP definition for each patient will be finalized before database lock and study unblinding.

The primary and secondary efficacy analyses will be based on the ITT population. Analyses based on the PP population will be considered supportive.

##### 14.4. Criteria for the Termination of the Trial

Criteria refers to the above reported statistical plan (section 14.2).

##### 14.5. Procedure for Accounting for Missing, Unused, and Spurious Data

All data will be checked for consistency and plausibility based on the nature of each collected variable before any statistical analysis. Data falling beyond limits of plausibility will be checked in the medical records and in the CRF to correct for possible typos or transcription errors. Changes in the database will be recorded together with date and name of the physician performing that correction and with the reason for correction.
Methods of multiple imputation will be considered if the missingness rate will be more than 20%.

##### 14.6. Inclusion in Analysis

All patients will be included in the analysis

##### 14.7. Procedures for Reporting any Deviation(s) from the Original Statistical Plan

All the statistical analysis procedures, all the possible deviations, and all the reporting operations will be performed according to the Statistical Principles for Clinical Trials (E9, ICH Harmonised Tripartite Guideline).
The statistical report will contain the log of any of the operation conducted on the database, plus, possible comments (preceded by a "%" symbol) if and when needed. Any deviation from the original statistical plan will be therefore reported in the log, together with the reason behind the deviation.

#### 15. DATA MANAGEMENT

##### 15.1. Source Data

Source documents are the first recorded data and from which participants' eCRF data are obtained. These include, but are not limited to, hospital records (from which medical history and previous and concurrent medication may be summarised into the eCRF), clinical and office charts, laboratory and pharmacy records, diaries, microfiches, radiographs, and correspondence.

All documents will be stored safely in confidential conditions. On all trial-specific documents, other than the signed consent, the participant will be referred to by the trial participant number/code, not by name.

##### 15.2. Access to Data

Direct access will be granted to authorised representatives from promoter and the regulatory authorities to permit trial-related monitoring, audits and inspections.

##### 15.3. Data Recording and Record Keeping

All trial data will be entered on an electronic CRFs.
The participants will be identified by a unique trial specific number and/or code in any database. The name and any other identifying detail will NOT be included in any trial data electronic file.

#### 16. QUALITY ASSURANCE PROCEDURES

The trial will be conducted in accordance to the current approved protocol, ICH GCP, relevant regulations and standard operating procedures.

#### 17. SERIOUS BREACHES

The Medicines for Human Use (Clinical Trials) Regulations contain a requirement for the notification of "serious breaches" to the MHRA within 7 days of the Promoter becoming aware of the breach.
A serious breach is defined as "A breach of GCP or the trial protocol which is likely to affect to a significant degree

(a) the safety or physical or mental integrity of the subjects of the trial; or

(b) the scientific value of the trial".

In the event that a serious breach is suspected the Promoter must be contacted within 1 working day. In collaboration with the C.I., the serious breach will be reviewed by the Sponsor and, if appropriate, the Sponsor will report it to the REC committee, Regulatory authority and the NHS host organisation within seven calendar days.

#### 18. ETHICAL AND REGULATORY CONSIDERATIONS

##### 18.1. Declaration of Helsinki

The Investigator will ensure that this trial is conducted in accordance with the principles of the Declaration of Helsinki.

##### 18.2. Guidelines for Good Clinical Practice

The Investigator will ensure that this study is conducted in accordance with relevant regulations and with Good Clinical Practice.

##### 18.3. Approvals

The protocol, informed consent form, participant information sheet and any proposed advertising material will be submitted to an appropriate Research Ethics Committee (REC) and regulatory authorities for written approval.

The Investigator will submit and, where necessary, obtain approval from the above parties for all substantial amendments to the original approved documents.

##### 18.4. Reporting

The CI shall submit once a year throughout the clinical trial, or on request, an Annual Progress Report to the REC. In addition, an End of Trial notification and final report will be submitted to the REC.

##### 18.5. Participant Confidentiality

The trial staff will ensure that the participants' anonymity is maintained. The participants will be identified only by initials and a participant ID number on the eCRF and any electronic database. All documents will be stored securely and only accessible by trial staff and authorised personnel. The trial will comply with the Data Protection Act, which requires data to be anonymised as soon as it is practical to do so.

##### 18.6. Expenses and Benefits

They will not be intended payments to participants and any other benefits.

##### 18.7. Other Ethical Considerations

Nothing to declare

#### 19. FINANCE AND INSURANCE

##### 19.1. Funding

None.

##### 19.2. Insurance

The study compares two accepted treatment modalities of patients with COVID-19. As per DM 17.12.2004, art 2, comma 4, patients will be covered by hospital insurance.
ASST Grande Ospedale Metropolitano will guarantee for patients'insurance.

#### 20. PUBLICATION POLICY

The final manuscript of the study will be prepared by the Principal Investigators.
The data will remain property of the Promoters.

### LAST PROTOCOL VERSION (VERSION 4.0)

**Enoxaparin for thromboprophylaxis in hospitalized COVID-19 patients: comparison of 40 mg o.d. versus 40 mg b.i.d. A randomized Clinical Trial**

**Internal Reference Number / Short title:** X-COVID 19

**Ethics Ref:**

**Date and Version No**: September 2020, Version 4.0

| | |
|---|---|
| **Chief Investigators:** | Marco Cattaneo, Dipartimento di Scienze della Salute, Università Degli Studi di Milano, Milan, Italy; Unità di Medicina 2, ASST Santi Paolo e Carlo, Milan, Italy;<br>Nuccia Morici, ASST Grande Ospedale Metropolitano Niguarda, Milan, Italy. |
| **Italian Investigators:** | Paolo Tarsia, Claudia Marini, Massimo Puoti, Luca Bonacchini (ASST Grande Ospedale Metropolitano Niguarda, Milan, Italy), Paolo Righini (IRCCS Policlinico San Donato, Milan, Italy), Gaetano Maria De Ferrari, Fabrizio D'Ascenzo (AOU Città della Salute e della Scienza, Presidio Molinette, Torino, Italy), Sergio Leonardi, Claudio Montalto (IRCCS Policlinico San Matteo, Pavia, Italy), Paolo Bonfanti (ASST Monza – Ospedale di Monza, Monza, Italy), Ilaria Battistoni, Matteo Francioni (AOU Ospedali Riuniti di Ancona, Ancona, Italy, Annamaria Municinò, Alessandro Rollero (ASL 3 Genovese, Genova, Italy), Giulio Stefanini, Antonio Voza (IRCCS Istituto Clinico Humanitas, Rozzano, Milan, Italy), Stefano Savonitto (ASST Lecco, Ospedale A. Manzoni, Lecco, Italy), Italo Porto (IRCCS Ospedale Policlinico San Martino, Genova), Francesca Cortellaro (Ospedale San Carlo Borromeo, ASST Santi Paolo e Carlo, Milano, Italy), Donato Bettega (Ospedale Sacra Famiglia, Provincia Lombardo-Veneta Fatebenefratelli, Erba, Como, Italy), Ruggero Giuliani (Casa Circondariale San Vittore, ASST-Santi Paolo e Carlo, Milano), Daniele Grosseto (Ospedale Infermi, Azienda USL Romagna, Rimini, Italy); Roberto Pedretti, Maria Francesca Barmina (IRCCS Multimedica, Sesto San Giovanni, Milano); Francesco Saverio Serino (ASL 4 Veneto, Covid Hospital, Jesolo, Italy), Massimo Antonio Di Pietro, Michele Trezzi (AUSL Toscana Centro, Ospedale San Jacopo). |
| **Russian Investigator** | Makatsariya Alexander (I.M. Sechenov First Moscow State Medical University), Nikita Lomakin (Central Clinical Hospital of the Presidential Department of Russian Federation) |
| **Sponsor:** | ASST Grande Ospedale Metropolitano Niguarda, Milan, Italy |
| **Funder:** | Italfarmaco |
| **Chief Investigator Signature:** | Marco Cattaneo<br><br>Nuccia Morici |
| **Statistician Signature:** | Laura Antolini<br><br>Maria Grazia Valsecchi |

**Confidentiality Statement:** This document contains confidential information that must not be disclosed to anyone other than the Sponsor, the Investigator Team, HRA, host organisation, and members of the Research Ethics Committee, unless authorised to do so.

### TABLE OF CONTENTS

1. KEY TRIAL CONTACTS 30
2. SYNOPSIS 31
3. ABBREVIATIONS 33
4. BACKGROUND AND RATIONALE 33
5. OBJECTIVES AND OUTCOME MEASURES 35
6. TRIAL DESIGN 36
7. PARTICIPANT IDENTIFICATION 37
   7.1 Trial participants 37
   7.2 Inclusion Criteria 37
   7.3 Exclusion Criteria 38
8 TRIAL PROCEDURES 38
   8.1 Recruitment 39
   8.2 Informed Consent 39
   8.3 Screening and Eligibility Assessment 39
   8.4 Randomisation, blinding and code-breaking 40
   8.5 Baseline Assessments 40
   8.6 Subsequent Visits 40
   8.7 Discontinuation/Withdrawal of Participants from Trial Treatment 41
   8.8 Definition of End of Trial 42
9. COMPLIANCE WITH TRIAL TREAMTENT 42
10. ACCOUNTABILITY OF TRIAL TREATMENT 42
11. CONCOMITANT MEDICATION 42
12. POST-TRIAL TREATMENT 42
13. SAFETY REPORTING 42
   13.1. Definitions 42
   13.2. Causality 43
   13.3. Procedures for Recording Adverse Events 43
   13.4. Reporting Procedures for Serious Adverse Events 44
   13.5. Expectedness 44
   13.6. SUSAR Reporting 44
   13.7. Safety Monitoring Committee 44
14. STATISTICS 45
   14.1. Description of Statistical Methods 45
   14.2. Number of Participants 45
   14.3. Populations Analysis 46
   14.4. Criteria for the Termination of the Trial 46
   14.5. Procedure for Accounting for Missing, Unused, and Spurious Data 46

14.6. Inclusion in Analysis 46
14.7. Procedures for Reporting any Deviation(s) from the Original Statistical Plan 46
15. DATA MANAGEMENT 47
15.1. Source Data 47
15.2. Access to Data 47
15.3. Data Recording and Record Keeping 47
16. QUALITY ASSURANCE PROCEDURES 47
17. SERIOUS BREACHES 47
18. ETHICAL AND REGULATORY CONSIDERATIONS 48
18.1. Declaration of Helsinki 48
18.2. Guidelines for Good Clinical Practice 48
18.3. Approvals 48
18.4. Reporting 48
18.5. Participant Confidentiality 48
18.6. Expenses and Benefits 49
19. FINANCE AND INSURANCE 49
19.1. Funding49
19.2. Insurance 49
20. PUBLICATION POLICY 49
21. REFERENCES 50

### 4. KEY TRIAL CONTACTS

| | |
|---|---|
| **Chief Investigator** | Marco Cattaneo, Dipartimento di Scienze della Salute, Università Degli Studi di Milano, Milan, Italy; Unità di Medicina 2, ASST Santi Paolo e Carlo, Milan, Italy. Phone 0250323095 - 0281844638; fax 0250323089;<br>Nuccia Morici, Unità di Cure Intensive Cardiologiche Dipartimento Cardio-toraco-vascolare, ASST Grande Ospedale Metropolitano Niguarda, Milan, Italy. Phone 0264442565, fax 0264442818, |
| **Sponsor** | ASST Grande Ospedale Metropolitano Niguarda, Milan, Italy |
| **Clinical Trials Unit** | Unità di Medicina, ASST Santi Paolo e Carlo, Milan, Italy / Malattie Infettive, ASST Grande Ospedale Metropolitano Niguarda, Milan, Italy |
| **Statisticians** | Laura Antolini, |
| **Committees** | **Comitato Etico Scientifico INMI Lazzaro Spallanzani (Ethical Committee)**<br>**Steering Committee**: Marco Cattaneo, Nuccia Morici, Enrico Ammirati, Marco Valgimigli, Stefano Savonitto. |

### 5. SYNOPSIS

| | |
|---|---|
| Trial Title | **Enoxaparin for thromboprophylaxis in hospitalized COVID-19 patients: comparison of 40 mg o.d. versus 40 mg b.i.d. A randomized Clinical Trial** |
| Internal ref. no. (or short title) | X-Covid 19 |
| Promoter | ASST Grande Ospedale Metropolitano Niguarda |
| Principal Investigator | Marco Cattaneo / Nuccia Morici |
| EudraCT Number | 2020-001708-41 |
| Clinical Phase | Phase IIIb |
| Trial Design | Open-label Multicentre Prospective Controlled Randomized Trial in patients with SARS-CoV-2 infection. Patients will be randomized 1:1 to 40 mg subcutaneous enoxaparin o.d. versus 40 mg enoxaparin b.i.d |
| Trial Participants<br>*Inclusion criteria* | • All-comers patients aged >=18 years and admitted to hospital with laboratory-confirmed SARS-CoV-2 infection. |
| *Exclusion criteria* | • Patients admitted directly to an intensive care unit;<br>• Estimated creatinine clearance <15 ml/min/1.73m$^2$;<br>• Patients needing anticoagulant for prior indication;<br>• Patients treated with heparin at any increased dose compared to prophylactic regimen before enrolment<br>• Patients at high bleeding risk or experiencing clinically significant bleeding;<br>• Patients involved in competitive clinical trials exploring antithrombotic treatments;<br>• Any other significant disease or disorder which, in the opinion of the investigator, may either put the participants at risk because of participation in the trial, or may influence the result of the trial, or the participant's ability to participate in the trial. |
| Planned Sample Size | A sample of 2712 subjects is required to test the superiority hypothesis with 80% power according to a two-sided 0.05 level of significance that 40 mg enoxaparin b.i.d. will decrease the primary endpoint compared to 40 mg enoxaparin o.d. An event rate of 10% was hypothesized in the control arm and 7% in the treatment arm, corresponding to a 33% relative risk reduction.<br>The trial will be designed with one interim analysis. This means that a preliminary test to potentially stop the trial for efficacy is performed after 492 out of 2712 planned subjects (246 randomized to enoxaparine 40 mg od and 246 randomized to enoxaparine 40 mg bid) have completed the trial. If the trial is not stopped early, a final test is performed once 100 percent of the planned subjects will be enrolled. For the interim analysis the proportion for the primary endpoint is estimated to be 0.07 in the half-dose anticoagulant group and 0.17 in the standard group. With a type I error $\alpha$ set to 0.05 and a power 1- $\beta$ set to 0.8, a total of 492 subjects will be required. |
| Treatment duration | In-hospital (at least 30 days of therapy) |

| | |
|---|---|
| Follow up duration | 30 days from the enrolment |
| Planned Trial Period | 13 months (Estimated end date May 2021) |
| Primary Objective | To compare the effects of 40 mg subcutaneous enoxaparin o.d. versus 40 mg enoxaparin b.i.d on the incidence of venous thromboembolism (VTE) [a composite of incident asymptomatic and symptomatic proximal deep vein thrombosis (DVT) diagnosed by serial compression ultrasonography (CUS), and symptomatic pulmonary embolism (PE) diagnosed by CT scan], in patients with SARS-CoV-2 infection. |
| Secondary Objectives | • To compare the effects of 40 mg subcutaneous enoxaparin o.d. versus 40 mg enoxaparin b.i.d on the incidence of in-hospital major complications, defined as the composite of death, VTE, use of mechanical ventilation, stroke, acute myocardial infarction and admission to an intensive care in patients with SARS-CoV-2.<br>• To compare each single component of the primary endpoint between the two groups.<br>• To compare maximum sequential organ failure assessment (SOFA) score between the two groups.<br>• To compare C-reactive protein, D-dimer, IL-6 and hs-troponin levels (as % above the upper reference limit [URL]) among the two groups.<br>• To compare the incidence of SARS-CoV-2-related Acute Respiratory Distress Syndrome (ARDS) between the two groups.<br>• To compare length of hospital stay between the two groups.<br>• To compare measures of right ventricular function at trans-thoracic echocardiography or CT between admission and follow-up, whenever available.<br>• To compare the incidence of 30 days adverse events, defined as composite of death, stroke and myocardial infarction |
| Safety Endpoints | • To compare the incidence of bleeding events according to the *ISTH bleeding scale* between the two groups.<br>• To compare the incidence of bleeding events according to *BARC classification* between the two groups. Only BARC 3 and 5 will be considered for secondary endpoint.<br>• To assess the incidence of heparin-induced thrombocytopenia in the two groups. |

#### 6. ABBREVIATIONS

| | |
|---|---|
| AE | Adverse event |
| AR | Adverse reaction |
| BARC | Bleeding Academic Research Consortium |
| CI | Chief Investigator |
| CRF | Case Report Form |
| CT | Clinical Trials |
| CTA | Clinical Trials Authorisation |
| CTRG | Clinical Trials and Research Governance |
| DMC/DMSC | Data Monitoring Committee / Data Monitoring and Safety Committee |
| DSUR | Development Safety Update Report |
| GCP | Good Clinical Practice |
| GP | General Practitioner |
| IB | Investigator's Brochure |
| ICF | Informed Consent Form |
| ICH | International Conference on Harmonisation |
| IMP | Investigational Medicinal Product |
| IRB | Independent Review Board |
| PI | Principal Investigator |
| PIL | Participant/ Patient Information Leaflet |
| REC | Research Ethics Committee |
| SAE | Serious Adverse Event |
| SAR | Serious Adverse Reaction |
| SDV | Source Data Verification |
| SMPC | Summary of Medicinal Product Characteristics |
| SOP | Standard Operating Procedure |
| SOFA | Sequential Organ Failure Assessment |
| SUSAR | Suspected Unexpected Serious Adverse Reactions |
| TMF | Trial Master File |

### 8. BACKGROUND AND RATIONALE

Critically ill medical patients are at heightened risk for venous thromboembolism (VTE) and, for this reason, they are treated with prophylactic doses of low molecular weight heparin (LMWH) (for instance, 40 mg enoxaparin). Patients with COVID-19, which is caused by SARS-CoV-2 infection, are apparently at particularly high risk of VTE, as a consequence of excessive activation of the hemostatic system which, in the most severe cases, could also be associated with the formation of microthrombi and clinically relevant disseminated intravascular coagulation.[1-4]

Based on personal communications by medical doctors working mostly in Northern Italy, there are concerns about the efficacy of thromboprophylaxis with low-molecular-weight heparin (LMWH) at the standard doses in these patients, given their apparently very high risk of thromboembolic complications. Therefore, in many Northern Italian hospitals these patients are given LMWH at much higher doses than those recommended for thromboprophylaxis: in some hospitals, full anticoagulant doses (100 U/Kg b.i.d.) are being used even in the absence of evidence of ongoing thromboembolic disease. This practice does not take into due account the obvious risk of major, potentially fatal bleeding that is associated with high dosing, in the absence of any evidence of higher efficacy of high doses compared to standard prophylactic doses.

An additional rationale for the use of high dose LMWH in many hospitals is that hyper-activation of the hemostatic system in COVID-19 patients could lead to the formation of microthrombi in pulmonary vessels[5], causing further respiratory distress, and in vessels of other vital organs, causing multi-organ failure (MOF). However, there is no demonstration that this therapeutic approach decreases the risk of microangiopathic thrombosis in these patients, as well as, for that matter, in other types of patients.

Given this scenario, we designed a randomized trial comparing standard prophylactic dose of subcutaneous enoxaparin (40 mg o.d.) with higher dose (40 mg b.i.d) with the aim of testing whether high-dose thromboprophylaxis is more effective than standard dose in preventing TEV in COVID-19 patients. This study would also allow us to test whether double-dose prophylaxis with enoxaparin would favourably affect the natural history of the disease, by decreasing the incidence of organ dysfunction associated with the formation of microthrombi in vital organs.

### 9. OBJECTIVES AND OUTCOME MEASURES

| Objectives | Outcome measures | Timepoint(s) of evaluation of these outcome measure (if applicable) |
|---|---|---|
| **Primary Objective**<br>• To compare the incidence of hospital-associated venous thromboembolism (HA-VTE) between the two groups (40 mg o.d. vs 40 mg b.i.d subcutaneous enoxaparin) | • ***Number (percentage) of patients who will experience*** the composite event.<br><br>• ***Relative risk*** will evaluate the association between the study outcome and treatment. | Discharge |
| **Secondary Objectives**<br>• To compare the incidence of in-hospital major complications, defined as the composite of death, HA-VTE, use of mechanical ventilation, major stroke, acute myocardial infarction and admission to an intensive care between the two groups.<br><br>• To compare each single component of the primary endpoint between the two groups.<br><br>• To compare maximum sequential organ failure assessment (SOFA) score between the two groups.<br><br>• To compare C-reactive protein, D-dimer, IL-6 and hs-troponin levels (as % above the upper reference limit [URL]) among the two groups.<br><br>• To compare the incidence of SARS-CoV-2- related Acute Respiratory Distress Syndrome (ARDS) between the two groups. | • ***Number (percentage) of patients who will experience*** the event. ***Relative risk*** will evaluate the association between the study outcome and treatment.<br><br>• ***Number (percentage) of patients who will experience*** the event. ***Relative risk*** will evaluate the association between the study outcome and treatment.<br><br>• Maximum sequential organ failure assessment (SOFA) score summarized by ***median and interquartile range (IQR) or mean and standard deviation*** according to variable distribution.<br><br>• Laboratory parameters will be indicated as ***median and interquartile range (IQR) or mean and standard deviation*** according to variable distribution.<br><br>• ***Number (percentage) of patients who will experience*** the event. ***Relative risk*** will evaluate the association between the study outcome and treatment. | Discharge<br><br>Discharge<br><br>Discharge<br><br>Discharge<br><br>Discharge<br><br>Discharge<br><br>Discharge<br><br>30 day |

| | | |
|---|---|---|
| • To compare length of hospital stay<br><br>• To compare measures of right ventricular function at trans-thoracic echocardiography or CT between admission and follow-up, whenever available.<br><br>• To compare the incidence of 30 days adverse events, defined as composite of death, stroke and myocardial infarction | • ***Median and interquartile range (IQR) or mean and standard deviation*** according to variable distribution.<br>• Echocardiographic parameters will be summarized by ***median and interquartile range (IQR) or mean and standard deviation*** according to variable distribution.<br><br>• ***Number (percentage) of patients who will experience*** the event. ***Relative risk*** will evaluate the association between the study outcome and treatment. | |
| **Safety endpoints**<br>• To assess the occurrence of bleeding events, according to the two classifications selected<br><br>• To assess the incidence of heparin-induced thrombocytopenia between the two groups. | • ***Number (percentage) of patients who will experience*** the event. ***Relative risk*** will evaluate the association between the study outcome and treatment.<br><br>• The incidence rate will be described across groups | During hospital stay<br><br>During hospital stay |

### 10. TRIAL DESIGN

Prospective controlled randomized, open-label multi-center trial.

Patients with COVID-19 (any hospitalized patients) with laboratory-confirmed SARS-CoV-2 infection will be randomized 1:1 to 40 mg enoxaparin/day subcutaneous versus 40 mg enoxaparin b.i.d. Standard dosage of Proton Pump Inhibitor (PPI) will be prescribed to each enrolled patient to reduce gastrointestinal (GI) bleeding risk. The chronic kidney disease epidemiology (CKD-EPI) formula will be used for eGFR calculation. Enoxaparin treatment will continue until discharge.

The study conduct will not change the usual clinical practice in terms of treatments and diagnostic procedures.

Patients will be monitored every 3 days with D-dimer, fibrinogen, complete blood count, lactate dehydrogenase (LDH), PT, APTT, C-reactive protein, procalcitonin only in the case of superinfection by a different agent, ferritin, creatine phosphokinase (CK), CK-MB, albumin, serum creatinine, aspartate transaminase (AST), alanine transaminase (ALT), high-sensitivity troponin, serum bilirubin and interleukine-6 along with other examinations according to clinical practice.

Patients will be monitored every seven days from the enrolment with CUS of the lower limbs for proximal DVT screening. For patients who display signs and symptoms of DVT, CUS will be performed as soon as possible in the same day. Proximal DVT, namely DVT of iliac, femoral and popliteal veins down to popliteal trifurcation, is defined by cross-sectional vein incompressibility or direct thrombus imaging with vein enlargement. Distal DVT might be reported if distal segments are explored.

For patients who display signs and symptoms of pulmonary embolism, a CT scan will be performed as soon as possible in the same day according to the indication of the attending physician.

Multidetector CT pulmonary angiography is the method of choice for imaging the pulmonary vasculature in patients with suspected pulmonary embolism, that is defined by contrast-filling defects.

With regard to secondary objective, the following definitions apply.

All-cause mortality includes all deaths, regardless of whether the cause of death is determined.

Cardiovascular death is defined as death due to atherosclerotic coronary heart disease, cerebrovascular accident or (complication of) peripheral embolization, and includes deaths due to acute MI, stroke, sudden death, non-sudden death, unwitnessed death, and procedure-related deaths. Non-cardiovascular death includes deaths due to all other causes.

Myocardial Infarction denotes the presence of acute myocardial injury detected by abnormal cardiac biomarkers [Cardiac troponin I (cTnI) and T (cTnT) increased above the 99th percentile upper reference limit (URL)] in the setting of evidence of acute myocardial ischaemia.

Stroke is defined as any new neurological deficit lasting >24 hours. CT or MRI are required to confirm the stroke and to distinguish between ischemic and hemorrhagic stroke. Strokes will be also classified as fatal or nonfatal: nonfatal stroke will be also classified as disabling and non disabling.

The total duration of the study is 13 months for enrolment, follow-up, data collection, cleaning and analysis.

### 11. PARTICIPANT IDENTIFICATION

#### 8.9 Trial participants

Patients directly admitted to hospital for suspected SARS-CoV-2 infection will be screened for randomization.

#### 8.10Inclusion Criteria

All the following inclusion criteria will be adopted:

All-comers patients aged >=18 years and admitted to hospital with laboratory-confirmed SARS-CoV-2 infection.

##### 8.11 Exclusion Criteria

The following patients will not be enrolled:

- Patients admitted directly to intensive care unit.
- Patients with estimated creatinine clearance <15 ml/min/1.73m$^2$.
- Patients needing anticoagulant for prior indication.
- Patients treated with heparin at any increased dose compared to prophylactic regimen before enrolment
- Patients at high bleeding risk or experiencing clinically significant bleeding;
- Patients involved in competitive clinical trials exploring antithrombotic treatments;
- Patients with any other significant disease or disorder which, in the opinion of the Investigator, may either put the participants at risk because of participation in the trial, or may influence the result of the trial, or the participant's ability to participate in the trial.

### 9 TRIAL PROCEDURES

Eligibility will be evaluated by the attending physician after the first visit and after confirmation of the diagnosis. The patient, properly informed about the study, will be able to decide to sign the informed consent and the privacy form. The physician will obtain a detailed anamnesis and he/she will carefully check the clinical parameters needed to define tissue perfusion (as detailed by the inclusion criteria) along with body temperature. The following laboratory data will be checked: haemochrome, renal function, serum electrolytes, liver function, coagulation, blood gas analysis. Laboratory examinations will be sequentially evaluated according to the flow chart timeline (section 8.6). If the patient develops a condition needing an anticoagulation, an increase in enoxaparin dosage to 100 UI/kg (1 mg/kg) b.i.d. might be considered if indicated and in the absence of contraindications. Enoxaparin treatments will continue until discharge. Serial CUS will be performed, the first on admission and then every seven days. Bleeding events will be registered and graded according to the International Society on Thrombosis and Haemostasis (ISTH) bleeding scale and Bleeding Academic Research Consortium (BARC) scale. Also BARC 2 bleedings will be recorded.

Pre-specified subgroup analyses will be performed for 1) D-dimer categories (>1 <2, 2, 3, 4, 5, 6 fold the ULN of the reference range); 2) estimated creatinine clearance (< versus >60 ml/min/1.73 m2); 3) severity of the respiratory symptoms (mild versus moderate-severe defined as respiratory rate ≥30 breaths/min; arterial oxygen saturation≤93% at rest; PaO2/FiO2 ≤300 mmHg); 4) concomitant antiplatelet therapy.

The identification of adverse reactions will be continuous during hospitalisation. The data will be recorded in an eCRF.

#### 9.1 Recruitment

Patients recruitment in Italy will be performed at the Cardiology Department, Malattie Infettive and Internal Medicine, ASST Grande Ospedale Metropolitano Niguarda, Milan, Italy; Medicina, Ospedale San Paolo, ASST Santi Paolo e Carlo, Milan, Italy; IRCCS Policlinico San Donato, Milan, Italy; AOU Città della Salute e della Scienza, Presidio Molinette, Torino, Italy; IRCCS Policlinico San Matteo, Pavia, Italy; ASST Monza – Ospedale di Monza, Monza, Italy; AOU Ospedali Riuniti di Ancona, Ancona, Italy; ASL 3 Genovese, Genova, Italy; IRCCS Istituto Clinico Humanitas, Rozzano, Milan, Italy; ASST Lecco, Ospedale A. Manzoni, Lecco, Italy; IRCCS Ospedale Policlinico San Martino, Genova, Italy; Ospedale San Carlo Borromeo, ASST Santi Paolo e Carlo, Milan, Italy; Ospedale Sacra Famiglia, Provincia Lombardo-Veneta Fatebenefratelli, Erba, Como, Italy; Casa Circondariale San Vittore, ASST-Santi Paolo e Carlo, Milano, Italy; Ospedale Infermi, Azienda USL Romagna, Rimini, Italy; IRCCS Multimedica, Sesto San Giovanni (Milano), Italy; ASL 4 Veneto, COVID Hospital, Jesolo (Venezia); AUSL Toscana Centro, Ospedale San Jacopo, Pistoia.

Patients recruitment in Russia will be performed at the I.M. Sechenov First Moscow State Medical University, Moscow, Russia; Central Clinical Hospital of the Presidential Department of Russian Federation, Moscow, Russia

#### 9.2 Informed Consent

The participant must personally sign and date the latest approved version of the Informed Consent form before any trial specific procedures.

Written and verbal versions of the Participant Information and Informed Consent will be presented to the participants detailing no less than: the exact nature of the trial; what it will involve the participant; the implications and constraints of the protocol; the known side effects and any risks involved in taking part. It will be clearly stated that the participant is free to withdraw from the trial at any time for any reason without prejudice to future care and with no obligation to give the reason for withdrawal.

The participant will be allowed as much time as wished, taking into consideration the urgency of treatments, to consider the information and the opportunity to ask the Investigator, his/her GP or other independent parties to decide whether he/she will participate in the trial. Written Informed Consent will then be obtained by means of participant dated signature and dated signature of the person who presented and obtained the Informed Consent. The person who obtained the consent must be suitably qualified and experienced, and authorized to do so by the Chief/Principal Investigator. The original signed Informed Consent will be given to the participant and a copy will be retained at the trial site.

#### 9.3 Screening and Eligibility Assessment

As clinically indicated, the screening procedures will be performed throughout demographics, medical history, concomitant medication, physical examination, laboratory tests and radiological findings (chest CT scans are encouraged over chest X-rays).
The screening procedure will be immediately followed by recruitment at arrival in the ward (patients will not be randomized in the emergency department), if allowed on basal evaluation and patient informed consent signature.

#### 9.4 Randomisation, blinding and code-breaking

This study will be a two-parallel group randomized, open-label, multi-center trial in hospitalized COVID-19 patients. Patients will be randomized by the attending physicians using an IVRS that randomizes at a ratio of 1:1 upon determination of eligibility.

A total sample of 2712 subjects is required to test the superiority hypothesis with 80% power according to a two-sided 0.5 level of significance that 40 mg enoxaparin subcutaneously bid/day (treatment arm) will decrease the primary endpoint compared to 40 mg enoxaparin/day (prophylaxis dose, that is considered the control arm as represents the standard of care for hospitalized patients that are bedridden). An absolute reduction of 3% is foreseen in the treatment arm (enoxaparin 40 mg b.i.d); an event rate of 10% was considered in the control arm (enoxaparin 40 mg o.d.) and 7% in the treatment arm.

The duration of the enrolment is 10 months from the obtaining of the approvals. The total duration of the study is 13 months for follow-up (1 months), data collection, cleaning and analysis (2 months). Randomisation will be done at the same visit as the baseline visit. There will not be a run-in period.

#### 9.5 Baseline Assessments

##### Basal evaluation (V1)

Every patient will be informed about the objectives of our study and he/she will be asked for consent to the trial. At basal visit and following ones, the physician will perform: physical examination, hemodynamic parameters and blood gas analysis check; CUS. Echocardiogram might be performed if requested by the attending physician.
The occurrence of organ dysfunction and failure, from admission through days spent in intensive care, will be assessed by the Sequential Organ Failure Assessment (SOFA) score.

#### 9.6 Subsequent Visits

After basal assessment, the following visits will be performed according to local clinical practice a check-up at discharge will be performed. Echocardiograms might be performed during hospitalization according to the indication of the attending physician. In case mechanical ventilation is required, ventilation modes and eventual variations of parameters will be recorded.

If subject is discharged before 30 days from enrolment, a telephonic follow up will be performed at 30 days to assess potential adverse events occurred during that period. A composite of overall mortality, myocardial infarction and stroke will be considered.

The schedule of clinical visits and assessment procedures is shown in the following flow chart.

**Flow chart**

| **Visit** | **V1** | **V2** | **V2** | **V3** | **V4** |
|---|---|---|---|---|---|
| **Time interval** | **Basal** | **To be checked each three days** | **To be checked each seven days** | **Discharge** | **30-day from the enrolment** |
| **Informed consent** | X | | | | |
| **Time from onset symptoms to enrolment** | X | | | | |
| **Physical examination** | X | | | | |
| **Height** | X | | | | |
| **Weight** | X | | | | |
| **Anamnesis** | X | | | | X |
| **BP (s/d/m)** | X | | | | |
| **HR** | X | X | | X | |
| **CUS** | X | | X | | |
| **Hb** | X | X | | X | |
| **Creatinine** | X | X | | X | |
| **Platelets** | X | X | | X | |
| **Leucocytes** | X | X | | X | |
| **INR** | X | X | | X | |
| **Bilirubin** | X | X | | X | |
| **AST** | X | X | | X | |
| **ALT** | X | X | | X | |
| **LDH** | X | X | | X | |
| **CRP** | X | X | | X | |
| **Troponin (HS)** | X | X | | X | |
| **IL-6** | X | X | | X | |
| **D-dimer** | X | X | | X | |
| **Fibrinogen** | X | X | | X | |
| **SOFA score** | X | X | | | |

#### 9.7 Discontinuation/Withdrawal of Participants from Trial Treatment

Each participant has the right to withdraw from the trial at any time. In addition, the Investigator may discontinue a participant from the trial at any time if the Investigator considers it necessary for any reason including:

- Ineligibility (either arising during the trial or retrospectively having been overlooked at screening)
- Significant protocol deviation
- Significant non-compliance with treatment regimen or trial requirements
- An adverse event which requires discontinuation of trial treatment or results in inability to continue to comply with trial procedures
- Disease progression which requires discontinuation of trial treatment or results in inability to continue to comply with trial procedures
- Withdrawal of Consent

Withdrawal from the trial will result in exclusion of the data for that participant from analysis
The reason for withdrawal will be recorded in the eCRF.
If the participant is withdrawn due to an adverse event, the Investigator will arrange for follow-up visits or telephone calls until the adverse event has resolved or stabilised.

##### 9.8 Definition of End of Trial

The end of trial is the date of the last visit/ telephone follow up/ home visit of the last participant.

#### 22. COMPLIANCE WITH TRIAL TREAMTENT

All patients will be treated in hospital ward, so compliance will be guaranteed by the investigators.

#### 23. ACCOUNTABILITY OF TRIAL TREATMENT

The study does will involve drugs usually administrated in clinical practice.

#### 24. CONCOMITANT MEDICATION

There are not any contraindicated medications. All used drugs, as per clinical indication, will be reported in the eCRF.

#### 25. POST-TRIAL TREATMENT

At the end of the study period, all surviving patients will be treated as per current clinical practice in the participating Centers.

#### 26. SAFETY REPORTING

##### 26.1. Definitions

| | |
|---|---|
| Adverse Event (AE) | Any untoward medical occurrence in a participant to whom a medicinal product or diagnostic-interventional procedure has been |

| | |
|---|---|
| | administered, including occurrences which are not necessarily caused by or related to that product or procedure. |
| Adverse Reaction (AR) | An untoward and unintended response in a participant to a medicinal product which is related to any dose administered to that participant.<br>The phrase "response to an investigational medicinal product" means that a causal relationship between a trial medication and an AE is at least a reasonable possibility, i.e. the relationship cannot be ruled out.<br>All cases judged by either the reporting medically qualified professional or the Sponsor as having a reasonable suspected causal relationship to the trial medication qualify as adverse reactions. |
| Serious Adverse Event (SAE) | A serious adverse event is any untoward medical occurrence that:<br>• Results in death<br>• Is life-threatening<br>• Requires inpatient hospitalisation or prolongation of existing hospitalisation<br>• Results in persistent or significant disability/incapacity<br>• Consists of a congenital anomaly or birth defect.<br>Other ‘important medical events’ may also be considered serious if they jeopardise the participant or require an intervention to prevent one of the above consequences.<br>NOTE: The term "life-threatening" in the definition of "serious" refers to an event in which the participant was at risk of death at the time of the event; it does not refer to an event which hypothetically might have caused death if it were more severe. |

NB: to avoid confusion or misunderstanding of the difference between the terms “serious” and “severe”, the following note of clarification is provided: “Severe” is often used to describe intensity of a specific event, which may be of relatively minor medical significance. “Seriousness” is the regulatory definition supplied above.

#### 26.2. Causality

The relationship of each adverse event to trial treatment must be determined by a medically qualified individual according to the following definitions:

**Related**: The adverse event follows a reasonable temporal sequence from trial treatment start. It cannot reasonably be attributed to any other cause.

**Not Related**: The adverse event is probably produced by the participant’s clinical state or by other modes of therapy administered to the participant.

#### 26.3. Procedures for Recording Adverse Events

All AEs occurring during the trial / or until the following two months that are observed by the Investigator or reported by the participant, will be recorded on the eCRF, whether or not attributed to trial treatment.

The following information will be recorded: description, date of onset and end date, severity, assessment of relatedness to trial treatment, other suspect drug or device and action taken. Follow-up information should be provided as necessary.
The severity of events will be assessed on the following scale: 1 = mild, 2 = moderate, 3 = severe.
AEs considered related to the trial treatment as judged by a medically qualified investigator will be followed either until resolution, or the event is considered stable.
It will be left to the Investigator's clinical judgment to decide whether an AE is of enough severity to require the participant's removal from treatment. A participant may also voluntarily withdraw from treatment due to what he or she perceives as an intolerable AE. If either of these occurs, the participant must undergo an end of trial assessment and be given appropriate care under medical supervision until symptoms cease, or the condition becomes stable.

#### 26.4. Reporting Procedures for Serious Adverse Events

All SAEs (other than those defined in the protocol as not requiring reporting) must be reported on the SAE reporting form to Eudravigilance within 24 hours of the Site Study Team becoming aware of the event.
Review of SAEs must be timely, taking into account the reporting time for a potential SUSAR.

#### 26.5. Expectedness

Expectedness will be determined according to the Investigators' Brochure/Summary of Product Characteristics.

#### 26.6. SUSAR Reporting

All SUSARs will be reported by the CI to the relevant Competent Authority and to the REC and other parties as applicable. For fatal and life-threatening SUSARS, this will be done no later than 7 calendar days after the investigator team is first aware of the reaction. Any additional information will be reported within 15 calendar days. Treatment codes will be un-blinded for specific participants.

#### 26.7. Safety Monitoring Committee

The aims of this committee include:

- To pick up any trends, such as increases in un/expected events, and take appropriate action
- To seek additional advice or information from investigators where required
- To evaluate the risk of the trial continuing and take appropriate action where necessary

### 27. STATISTICS

#### 27.1. Description of Statistical Methods

All protocol-defined endpoints will be measured from the time of randomization. Descriptive statistics of baseline characteristics of patients randomized to the two groups will be reported. Continuous variables will be summarized by means, standard deviations, medians, inter-quartile ranges and minimum and maximum values. Categorical variables will be summarized by frequencies and percentages.

Data will be analysed according to the intention-to-treat principle. In addition, a per-protocol analysis of the primary endpoint, considering patients without major protocol violations (i.e. patients who crossed over), will be performed to evaluate the robustness of the data.

A test for difference of proportions will be carried out to compare in-hospital composite events among patients randomized to the two groups. A relative risk will be calculated to quantify the magnitude of the half anticoagulant effect.

For what regards continuous secondary endpoints, maximum SOFA score will be compared among the study groups through the t-test or Wilcoxon rank-sum test, according to the distribution of study endpoints. The same approach will be applied for laboratory examinations and times included in the secondary endpoints.

All tests will be two-sided and a p-value of 0.05 or less will be considered statistically significant. No adjustments for multiple testing will be applied in the analysis of secondary endpoints and in the subgroup analysis, therefore, inferences drawn from the reported p-values and 95% CIs will be considered exploratory. If a p-value is less than 0.001 it will be reported as "<0.001".

#### 27.2. Number of Participants

**Summary Statements**

The proportion for the primary endpoint is estimated to be 0.07 in the half-dose anticoagulant dose and 0.10 in the standard group. With a type I error α set to 0.05 and a power 1- β set to 0.8, a total of 2712 subjects will be enrolled in the trial. The trial will be designed with one interim analysis. This means that a preliminary test to potentially stop the trial for efficacy is performed after 492 out of 2712 planned subjects (246 randomized to enoxaparine 40 mg od and 246 randomized to enoxaparine 40 mg bid) have completed the trial. If the trial is not stopped early, a final test is performed once 100 percent of the planned subjects will be enrolled. For the interim analysis the proportion for the primary endpoint is estimated to be 0.07 in the half-dose anticoagulant group and 0.17 in the standard group. With a type I error α set to 0.05 and a power 1- β set to 0.8, a total of 492 subjects will be required.

#### 27.3. Populations Analysis

For this trial, the following populations will be defined and used in the analysis and/or presentation of the data.

**Safety population:** The safety population will be classified according to the actual treatment on enoxaparin 40 mg b.i.d. or control, received.

**Intent-to-treat (ITT) population***:* Treatment classification for ITT analysis will be based on the randomized treatment.

**Per-Protocol (PP) population:** The PP population will be defined as all ITT patients who received assigned to enoxaparin 40 mg b.i.d. and will be without major protocol violations. Specifically, subjects may be excluded from PP population if any of the following criteria are met:

- Inclusion/Exclusion criteria violations;
-

The PP definition for each patient will be finalized before database lock and study unblinding.

The primary and secondary efficacy analyses will be based on the ITT population. Analyses based on the PP population will be considered supportive.

#### 27.4. Criteria for the Termination of the Trial

Criteria refers to the above reported statistical plan (section 14.2).

#### 27.5. Procedure for Accounting for Missing, Unused, and Spurious Data

All data will be checked for consistency and plausibility based on the nature of each collected variable before any statistical analysis. Data falling beyond limits of plausibility will be checked in the medical records and in the CRF to correct for possible typos or transcription errors. Changes in the database will be recorded together with date and name of the physician performing that correction and with the reason for correction.
Methods of multiple imputation will be considered if the missingness rate will be more than 20%.

#### 27.6. Inclusion in Analysis

All patients will be included in the analysis

#### 27.7. Procedures for Reporting any Deviation(s) from the Original Statistical Plan

All the statistical analysis procedures, all the possible deviations, and all the reporting operations will be performed according to the Statistical Principles for Clinical Trials (E9, ICH Harmonised Tripartite Guideline).

The statistical report will contain the log of any of the operation conducted on the database, plus, possible comments (preceded by a "%" symbol) if and when needed. Any deviation from the original statistical plan will be therefore reported in the log, together with the reason behind the deviation.

### 28. DATA MANAGEMENT

#### 28.1. Source Data

Source documents are the first recorded data and from which participants' eCRF data are obtained. These include, but are not limited to, hospital records (from which medical history and previous and concurrent medication may be summarised into the eCRF), clinical and office charts, laboratory and pharmacy records, diaries, microfiches, radiographs, and correspondence.

All documents will be stored safely in confidential conditions. On all trial-specific documents, other than the signed consent, the participant will be referred to by the trial participant number/code, not by name.

#### 28.2. Access to Data

Direct access will be granted to authorised representatives from promoter and the regulatory authorities to permit trial-related monitoring, audits and inspections.

#### 28.3. Data Recording and Record Keeping

All trial data will be entered on an electronic CRFs.

The participants will be identified by a unique trial specific number and/or code in any database. The name and any other identifying detail will NOT be included in any trial data electronic file.

### 29. QUALITY ASSURANCE PROCEDURES

The trial will be conducted in accordance to the current approved protocol, ICH GCP, relevant regulations and standard operating procedures.

### 30. SERIOUS BREACHES

The Medicines for Human Use (Clinical Trials) Regulations contain a requirement for the notification of "serious breaches" to the MHRA within 7 days of the Promoter becoming aware of the breach.
A serious breach is defined as “A breach of GCP or the trial protocol which is likely to affect to a significant degree

(a) the safety or physical or mental integrity of the subjects of the trial; or

(b) the scientific value of the trial”.

In the event that a serious breach is suspected the Promoter must be contacted within 1 working day. In collaboration with the C.I., the serious breach will be reviewed by the Sponsor and, if appropriate, the Sponsor will report it to the REC committee, Regulatory authority and the NHS host organisation within seven calendar days.

### 31. ETHICAL AND REGULATORY CONSIDERATIONS

#### 31.1. Declaration of Helsinki

The Investigator will ensure that this trial is conducted in accordance with the principles of the Declaration of Helsinki.

#### 31.2. Guidelines for Good Clinical Practice

The Investigator will ensure that this study is conducted in accordance with relevant regulations and with Good Clinical Practice.

#### 31.3. Approvals

The protocol, informed consent form, participant information sheet and any proposed advertising material will be submitted to an appropriate Research Ethics Committee (REC) and regulatory authorities for written approval.
The Investigator will submit and, where necessary, obtain approval from the above parties for all substantial amendments to the original approved documents.

#### 31.4. Reporting

The CI shall submit once a year throughout the clinical trial, or on request, an Annual Progress Report to the REC. In addition, an End of Trial notification and final report will be submitted to the REC.

#### 31.5. Participant Confidentiality

The trial staff will ensure that the participants’ anonymity is maintained. The participants will be identified only by initials and a participant ID number on the eCRF and any electronic database. All documents will be stored securely and only accessible by trial staff and authorised personnel. The trial will comply with the Data Protection Act, which requires data to be anonymised as soon as it is practical to do so.

##### 31.6. Expenses and Benefits

They will not be intended payments to participants and any other benefits.

##### 31.7. Other Ethical Considerations

Nothing to declare

#### 32. FINANCE AND INSURANCE

##### 32.1. Funding

Italfarmaco will provide funding for clinical study.

##### 32.2. Insurance

The study compares two accepted treatment modalities of patients with COVID-19.
As per DL 8 aprile 2020, n. 23, art 40, comma 5, “Disposizioni urgenti materia di sperimentazione dei medicinali per l’emergenza epidemiologica da COVID”, for COVID-related No-Profit studies, no insurance is needed in Italy.
For Russian centers, no insurance is needed per local laws.

#### 33. PUBLICATION POLICY

The final manuscript of the study will be prepared by the Principal Investigators.
The data will remain property of the Promoters.

### SUMMARY OF MAIN PROTOCOL CHANGES

The original version of the protocol was 1.0 and was dated April 2020. Below the rationale and the summary of changes introduced with the various amendments.

##### *Amendment 1 to the protocol – Version 2.0 – May 2020:*

**Amendment Rationale:**

- Updated the primary endpoint:
  Primary endpoint was misleading, missing the specification that only proximal deep vein thrombosis has to be considered in the composite analysis

- Changes to trial design:
  Removed the need to randomize within 12 hours of hospital admission to allow for better recruitments.

- Modified exclusion criteria:
  Modified the criteria for subjects participating in other clinical trials to exclude only those enrolled in competitive clinical trials with antithrombotic agents; all subjects with higher than prophylactic doses of heparin were also excluded.

- Added the interim analysis:
  To allow an early evaluation of trial efficacy and safety

- Updated investigator list and recruiting centers
  To allow an increased rate of enrollment by adding new centers to the trial

**Summary of changes:**

- Updated the primary objective (section 2)

- Modification to trial design (section 6)

- Modified the exclusion criteria (section 7.3)

- Updated the list of participating centers (section 8.1)

- Added an ad-interim analysis (section 14.2)

##### *Amendment 2 to the protocol – Version 3.0 – June 2020:*

**Amendment Rationale:**

- The trial was modified in order to become an international multicenter trial:
  Change was proposed to permit a better enrollment since Russia was undergoing a high incidence of infections

- Added the references to the sponsor allowing the internationalization of the study:

Sponsor’s funding was needed to allow opening centers in Russia by stipulating collaborations with local CRO

- Updates on the insurance policies:
  Russians’ centers required the stipulation of a local insurance policy

- Updated investigator list and recruiting centers
  To allow an increased rate of enrollment by adding new centers to the trial

**Summary of changes:**

- Modified the recruitment to include Russian centers (section 8.1)
- Updated the list of participating centers (section 8.1)
- Updated the funding references (Heading and section 19.1)
- Updated the insurance policies (section 19.2)

##### *Amendment 3 of the protocol – Version 4.0 – September 2020:*

**Amendment Rationale:**

- Modified trial design:
  Updated wordings for enoxaparin treatments that were ambiguous: added the specification that treatment will continue until discharge

- Added a secondary endpoint:
  “To compare the incidence of 30 days adverse events, defined as composite of death, stroke and myocardial infarction” , considering the increasing reported association between COVID disease and cardiovascular events.

- Modified the endpoint evaluation timepoints:
  Timepoints were modified from a 30-day time to Discharge to cover the usual period managed with thromboprophylaxis in medical patients.

- Increased the duration of the study to 13 months:
  Change was needed due to slow enrollment rates

- Updated information on Follow Up procedures:
  The follow up procedures were better described, specifying that a telephonic 30-day follow up evaluation is needed for subjects discharged before that timepoint

- Updates on the insurance policies:
  The need for an insurance was not needed in Russia for COVID related studies, thus it was removed

- Updated investigator list and recruiting centers

To allow an increased rate of enrollment by adding new centers to the trial

**Summary of changes:**

- Added a new secondary endpoint (section 2)
- Modified endpoints timepoints (section 5)
- Updated trial design (section 6 and section 8)
- Updated the list of participating centres (section 8.1)
- Modified the end study date and duration (section 6 and section 8.4)
- Clarified follow-up procedures (section 8.6)
- Updated the insurance policies (section 19.2)

### COMPLETE LIST OF STUDY OUTCOMES

**Primary Objective:**

- To compare the effects of 40 mg subcutaneous enoxaparin o.d. versus 40 mg enoxaparin b.i.d. on the incidence of venous thromboembolism (VTE) (a composite of incident asymptomatic and symptomatic proximal deep vein thrombosis (DVT) diagnosed by serial compression ultrasonography (CUS), and symptomatic pulmonary embolism (PE) diagnosed by CT scan), in patients with SARS-CoV-2 infection.

**Secondary Objectives:**

- To compare the effects of 40 mg subcutaneous enoxaparin o.d. versus 40 mg enoxaparin b.i.d. on the incidence of in-hospital major complications, defined as the composite of death, VTE, use of mechanical ventilation, stroke, acute myocardial infarction and admission to an intensive care in patients with SARS-CoV-2.
- To compare each single component of the primary endpoint between the two groups.
- To compare maximum sequential organ failure assessment (SOFA) score between the two groups.
- To compare C-Reactive Protein, D-dimer, IL-6 and HS-troponin levels (as % above the upper reference limit [URL]) among the two groups.
- To compare the incidence of SARS-CoV-2-related acute respiratory distress syndrome (ARDS) between the two groups.
- To compare length of hospital, stay between the two groups.
- To compare measures of right ventricular function at trans-thoracic echocardiography or CT between admission and follow-up, whenever available.
- To compare the incidence of 30 days adverse events, defined as composite of death, stroke and myocardial infarction.

**Safety Endpoints:**

- To compare the incidence of bleeding events according to the *ISTH bleeding scale* between the two groups.
- To compare the incidence of bleeding events according to *BARC classification* between the two groups. Only BARC 3 and 5 will be considered for secondary endpoint.
- To assess the incidence of heparin-induced thrombocytopenia in the two groups.

#### Definitions of Outcome Events

- **All-cause death:** Death due to any causes in the first 30 days. The steering committee made the decision not to adjudicate the cause of death whenever inaccuracies were possible and no systematic autopsies were performed.

- **Deep venous thrombosis**: Incident symptomatic and asymptomatic proximal deep vein thrombosis diagnosed in a major vein of the legs using compression ultrasound.

- **Pulmonary embolism:** Any pulmonary embolism found at autopsy or diagnosed on echocardiography, V/Q scan, CT angiography or invasive pulmonary angiography. ASH 2020 Guidelines were followed for management.(1)

- **Ischemic stroke:** An episode of neurological dysfunction caused by focal cerebral, spinal, or retinal infarction.(2)

- **Type 1 myocardial infarction (T1MI)**: It was defined as: "An acute myocardial injury with clinical evidence of acute myocardial ischaemia and with detection of a rise and/or fall of cTn values with at least one value above the 99th percentile URL and at least one of the following: Symptoms of myocardial ischaemia; New ischaemic ECG changes; Development of pathological Q waves; Imaging evidence of new loss of viable myocardium or new regional wall motion abnormality in a pattern consistent with an ischaemic aetiology; Identification of a coronary thrombus by angiography or autopsy (not for types 2 or 3 MIs)"(3)

- **ARDS:** An acute diffuse, inflammatory lung injury, leading to increased pulmonary vascular permeability, increased lung weight, and loss of aerated lung tissue. It was defined, following Berlin 2012 definition, as the acute onset of hypoxemia with bilateral opacities on chest imaging, with no evidence of left atrial hypertension.(4)

- **Heparin-induced thrombocytopenia**: An adverse drug reaction mediated by platelet-activating antibodies that target complexes of platelet factor 4 and heparin. Diagnosed following ASH 2018 Guidelines (5)

- **BARC Scale** (6)
  Type 0: no bleeding
  Type 1: bleeding that is not actionable and does not cause the patient to seek unscheduled performance of studies, hospitalization, or treatment by a healthcare professional; may include episodes leading to self-discontinuation of medical therapy by the patient without consulting a healthcare professional.
  Type 2: any overt, actionable sign of hemorrhage (eg, more bleeding than would be expected for a clinical circumstance, including bleeding found by imaging alone) that does not fit the criteria for type 3, 4, or 5 but does meet at least one of the following criteria: (1) requiring nonsurgical, medical intervention by a healthcare professional, (2) leading to hospitalization or increased level of care, or (3) prompting evaluation.
  Type 3a: Overt bleeding plus hemoglobin drop of 3 to <5 g/dL (provided hemoglobin drop is related to bleed); Any transfusion with overt bleeding.
  Type 3b: Overt bleeding plus hemoglobin drop ≥5 g/dL (provided hemoglobin drop is related to bleed); Cardiac tamponade; Bleeding requiring surgical intervention for control (excluding dental/nasal/skin/hemorrhoid); Bleeding requiring intravenous vasoactive agents.

Type 3c: Intracranial hemorrhage (does not include microbleeds or hemorrhagic transformation, does include intraspinal); Subcategories confirmed by autopsy or imaging or lumbar puncture; Intraocular bleed compromising vision.
Type 4: CABG-related bleeding: Perioperative intracranial bleeding within 48 h; Reoperation after closure of sternotomy for the purpose of controlling bleeding; Transfusion of ≥5 U whole blood or packed red blood cells within a 48-h period; Chest tube output ≥2L within a 24-h period.
Type 5: fatal bleeding
Type 5a: Probable fatal bleeding; no autopsy or imaging confirmation but clinically suspicious
Type 5b: Definite fatal bleeding; overt bleeding or autopsy or imaging confirmation

- **ISTH definition of major bleeding in non-surgical patients:**
  Fatal bleeding, and/or
  Symptomatic bleeding in a critical area or organ, such as intracranial, intraspinal, intraocular, retroperitoneal, intra-articular or pericardial, or intramuscular with compartment syndrome, and/or
  Bleeding causing a fall in haemoglobin level of 20 g $L^{-1}$ (1.24 mmol $L^{-1}$) or more, or leading to transfusion of two or more units of whole blood or red cells. (7)

- **ISTH definition of major bleeding in surgical patients:**
  Fatal bleeding and/or
  Bleeding that is symptomatic and occurs in a critical area or organ, such as intracranial, intraspinal, intraocular, retroperitoneal, pericardial, in a non-operated joint, or intramuscular with compartment syndrome, assessed in consultation with the surgeon and/or
  Extrasurgical site bleeding causing a fall in haemoglobin level of 2 g/dL (1.24 mmol/L) or more, or leading to transfusion of two or more units of whole blood or red cells, with temporal association within 24–48 h to the bleeding, and/or
  Surgical site bleeding that requires a second intervention (open, arthroscopic, endovascular) or a hemarthrosis of sufficient size as to interfere with rehabilitation by delaying mobilization or delayed wound healing, resulting in prolonged hospitalization or a deep wound infection, and/or
  Surgical site bleeding that is unexpected and prolonged and/ or sufficiently large to cause hemodynamic instability, as assessed by the surgeon. There should be an associate fall in haemoglobin level of at least 2 g/dL (1.24 mmol/L), or transfusion, indicated by the bleeding, of at least two units of whole blood or red cells, with temporal association within 24 h to the bleeding.
  The period for collection of these data is from start of surgery until five half-lives after the last dose of the drug with the longest half-life and with the longest treatment period (in case of unequal active treatment durations).
  The population is those who have received at least one dose of the study drug.(8)

- **SOFA Score:** The Sequential Organ Failure Assessment Score, evaluating organ disfunction in critically ill patients.

Respiration (P/F), Coagulation (Platelets), Liver (Bilirubin), Cardiovascular (hypotension and amine use) , Central Nervous System (Glasgow Coma Scale) and Renal (Creatinine or urine output) were assessed.(9,10)

| **Variables/score** | **0** | **1** | **2** | **3** | **4** |
|---|---|---|---|---|---|
| **$PaO_2/FiO_2$ (mmHg)** | >400 | ≤400 | ≤300 | ≤200 | ≤100 |
| **Platelets ($x10^3/\mu l$)** | >150 | ≤150 | ≤100 | ≤50 | ≤20 |
| **Bilirubin (mg/dl)** | <1.2 | 1.2-1.9 | 2-5.9 | 6-11.9 | >12 |
| **Cardiovascular (Hg/kg/min)** | - | MAP<70 | Dop≤5 | Dop>5 (Epi≤0.1) | Epi>0.1 |
| **Glasgow Coma Scale** | 15 | 13-14 | 10-12 | 6-9 | <6 |
| **Creatinine (mg/dl)** | <1.2 | 1.2-1.9 | 2-3.4 | 3.5-4.9 | >5 |

MAP, Mean Arterial Pressure; Dop, Dopamine; Epi, Epinephrine.

- **Borg Scale:** (11) a 6-to-20-grade scale for ratings of perceived exertion.
  6: No exertion at all
  7: Very, very light
  9: Very light
  11: Fairly light
  13: Somewhat hard
  15: Hard
  17: Very hard
  19: Very, very hard

### CTPA COMPARISON BETWEEN GROUPS

Number of total TC, with or without contrast, in the different centers, divided by treatment arm

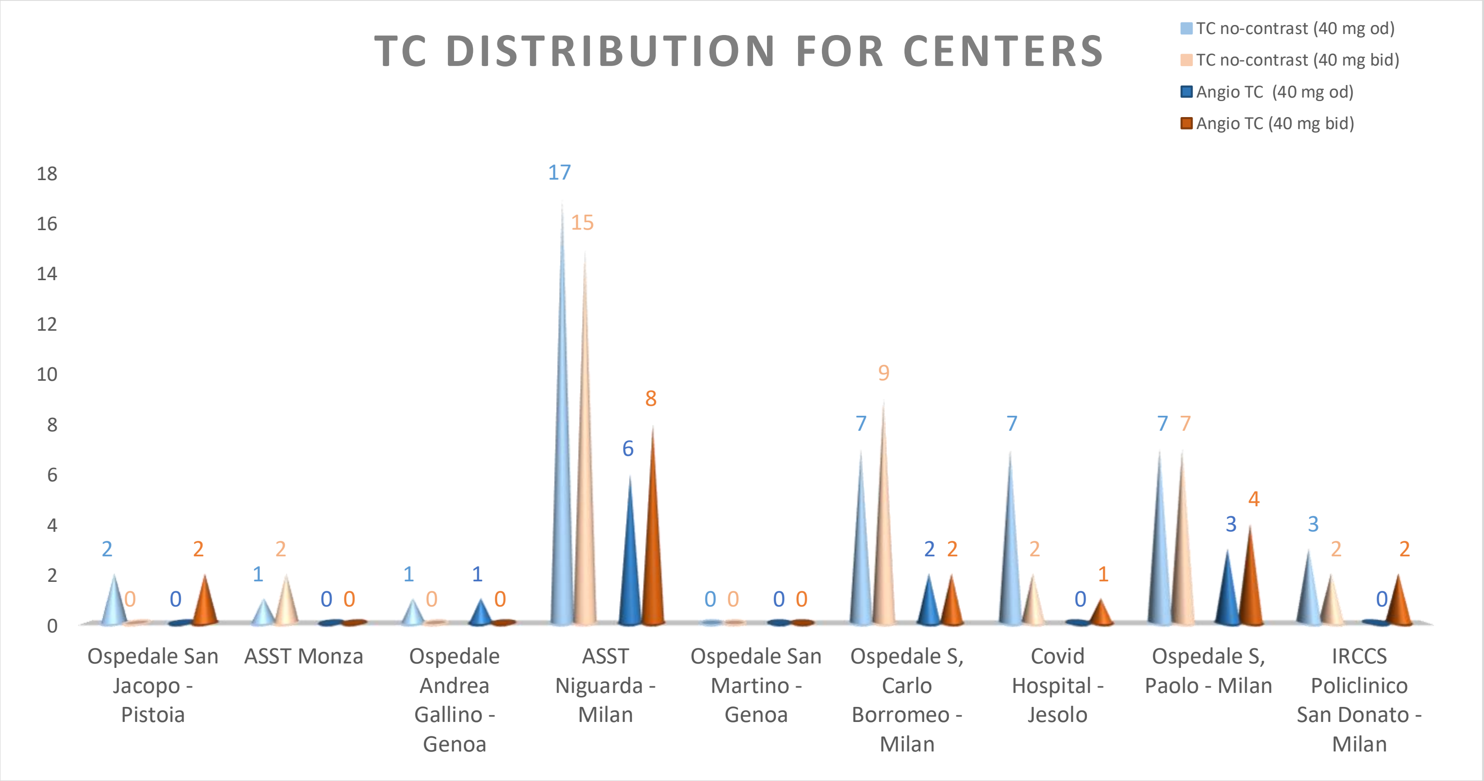

Symptoms for requiring Angio TC examination (multiple symptoms may have warranted for requiring examination)

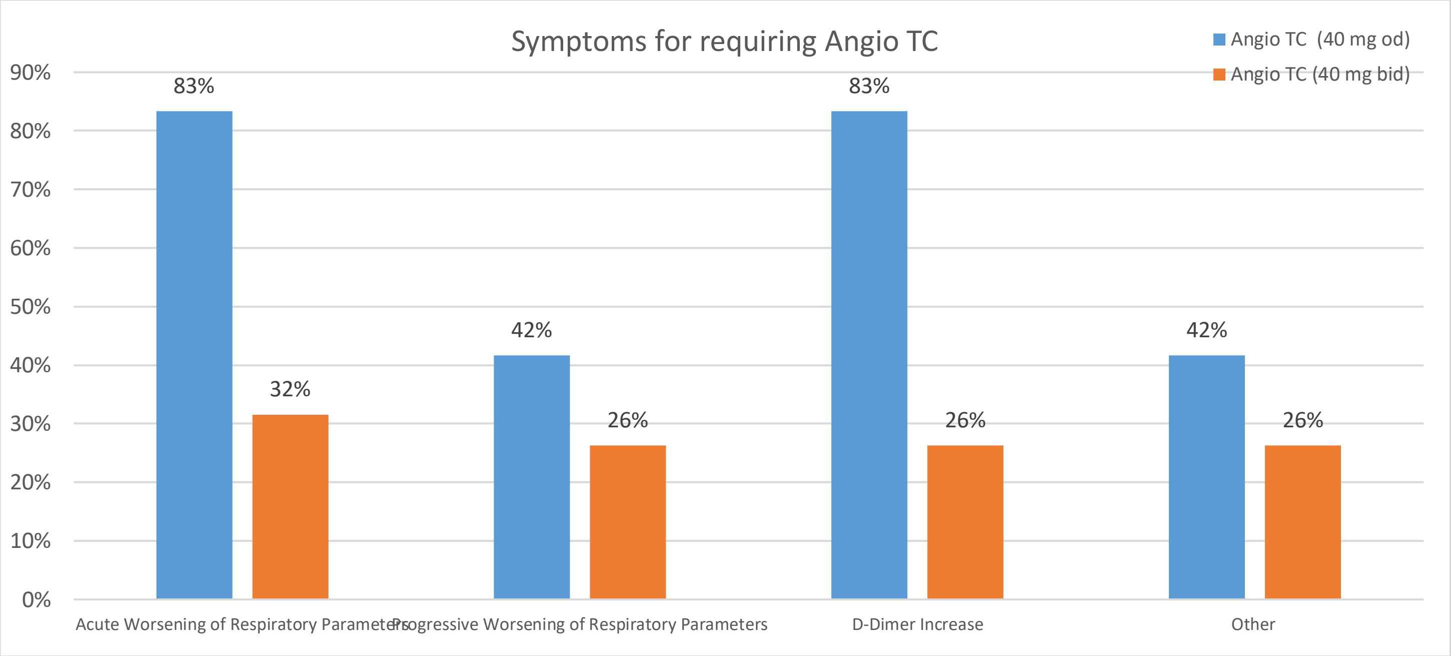

### CHARACTERISTICS OF SUBJECT WITH PULMUNARY EMBOLISM

The following table summarizes the clinical characteristics, treatments and symptoms of subjects that underwent pulmonary embolism.

| | | 001 | 002 | 003 | 004 | 005 | 006 |
|---|---|---|---|---|---|---|---|
| ARM | ***Treatment*** | 40 mg od | 40 mg od | 40 mg od | 40 mg od | 40 mg od | 40 mg od |
| ANAGRAPHICS | ***Sex*** | M | M | M | M | M | M |
| | ***Ethnicity*** | CAUCASIC | CAUCASIC | CAUCASIC | CAUCASIC | ARAB | CAUCASIC |
| SYMPTOMS | ***Fever*** | Yes | No | Yes | No | Yes | No |
| | ***Mialgia*** | No | No | Yes | Yes | No | No |
| | ***Asthenia*** | No | No | Yes | No | No | No |
| | ***Headache*** | No | No | No | No | No | No |
| | ***Asthma*** | No | No | Yes | No | No | No |
| | ***Cough*** | No | No | Yes | Yes | Yes | Yes |
| | ***Dyspnea*** | No | No | Yes | Yes | No | Yes |
| | ***Nausea/ Vomiting*** | No | No | No | No | No | No |
| | ***Diarrhea*** | No | No | No | No | No | No |
| | ***Other Symptoms*** | No | No | No | No | Epigastralgia | Chest Pain, Cardiopalmus |
| EXAMS | ***Pulmunary Examination*** | | | VM reduction | normal | nds | respiratory insufficiency, mild intercostal algia |
| | ***TC Evaluation*** | ground-glass bilateral multiple thickening | | negative for pulmonary embolism | bilateral ground-glass and right basal thickening with right pleuric effusion | Polmonite bilaterale AngioTC positiva per EP | Apical centrilobular emphysema, global increase in lung parenchyma density from confluent subsolid densities prevalent in both upper lobes, Corads 5, parenchymal commitment estimated about 30-40% |
| BASELINE THERAPY | ***Antiaggregants*** | Acetylsalicilic Acid 100mg bid | | Acetylsalicilic Acid 100mg bid | | | |
| | ***Anticoagulants*** | | | | | | |
| | ***Beta-Blockers*** | Nebivolol 1,25 mg sid | | Atenolol 7,5 mg bid | | | |

| | | | | | | | |
|---|---|---|---|---|---|---|---|
| | ***Calcium Antagonists*** | | | | | | |
| | ***Ace Inhibitors*** | Ramipril 10 mg sid | | Elanapril 10 mg bid | | | |
| | ***Sartans*** | | | | | | Olmesartan Medoxomil 10 mg od |
| | ***Sacubitril / Valsartan*** | | | | | | |
| | ***Diuretics*** | | | | | | Hydrochlorothiazide |
| | ***MRAs*** | | | | | Dexamethasone 6 mg od | |
| | ***Ivabradin*** | | | | | | |
| | ***Antiarrhytmics*** | | | | | | |
| | ***Statins*** | | | Rosuvastatin 40 mg od | | | |
| | ***Oral Hypoglicemics*** | | | | | Xelevia 100 mg od | |
| | ***Insulin*** | | | | | Yes | |
| | ***Immuno Suppressants*** | | | Dexamethasone 6 mg od | | | |
| | ***Antivirals*** | | | Remdesevir 100 mg od | | | |
| | ***Broncodilators*** | | | | | | Aclidinium Bromide |
| IN-HOSPITAL THERAPY | ***Antibiotics*** | | | | | | |
| | ***Steroids*** | | | | | | Dexamethasone 1.5 ml od |
| | ***Antithrombotics*** | | | | | | |
| | ***Gastric Protector*** | | | | | | Omeprazol 20 mg od |
| | ***Antiarrhytmics*** | | | | | | |
| | ***Inotropes*** | | | | | | |
| | ***Vasopressors*** | | | | | | |
| | ***Other Drugs*** | | | | | | Omelsartan Medoxomil 10 mg od |
| FU | ***Follow Up Status*** | Alive | Lost to FU | Alive | Alive | Alive | Alive |

### ENROLLMENTS INFORMATIONS

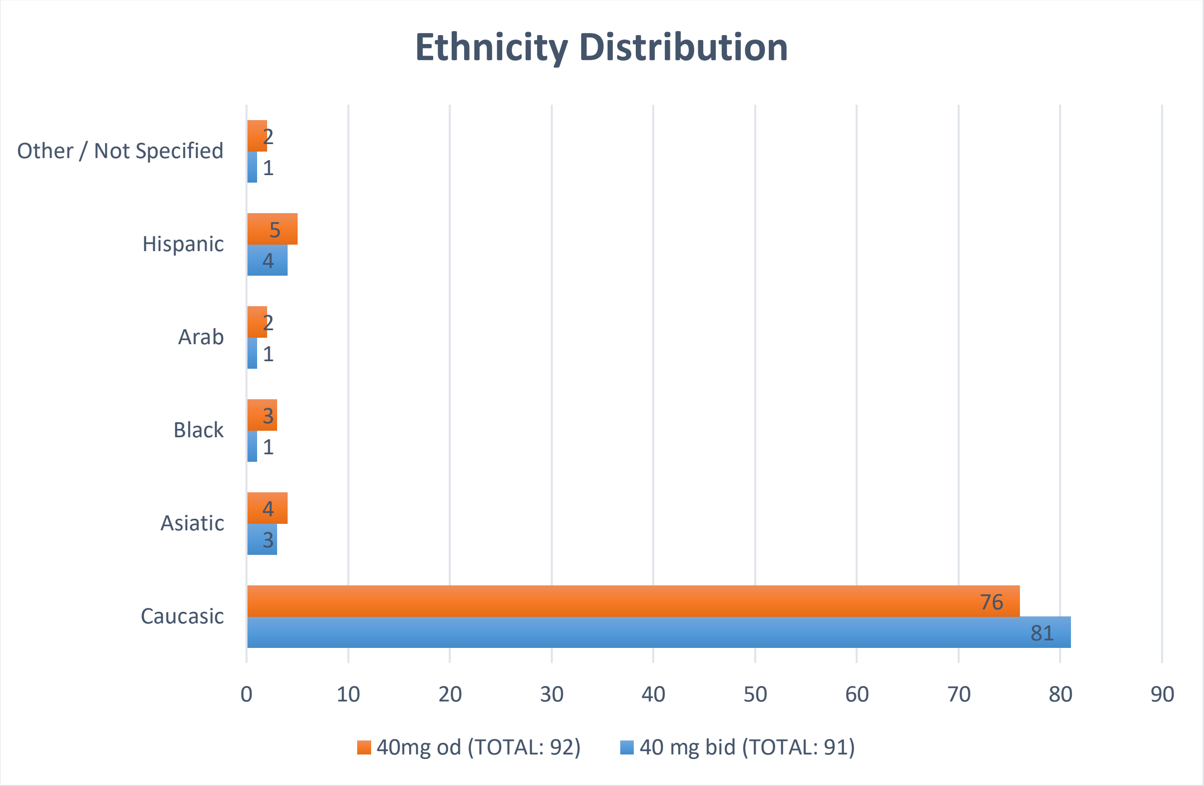

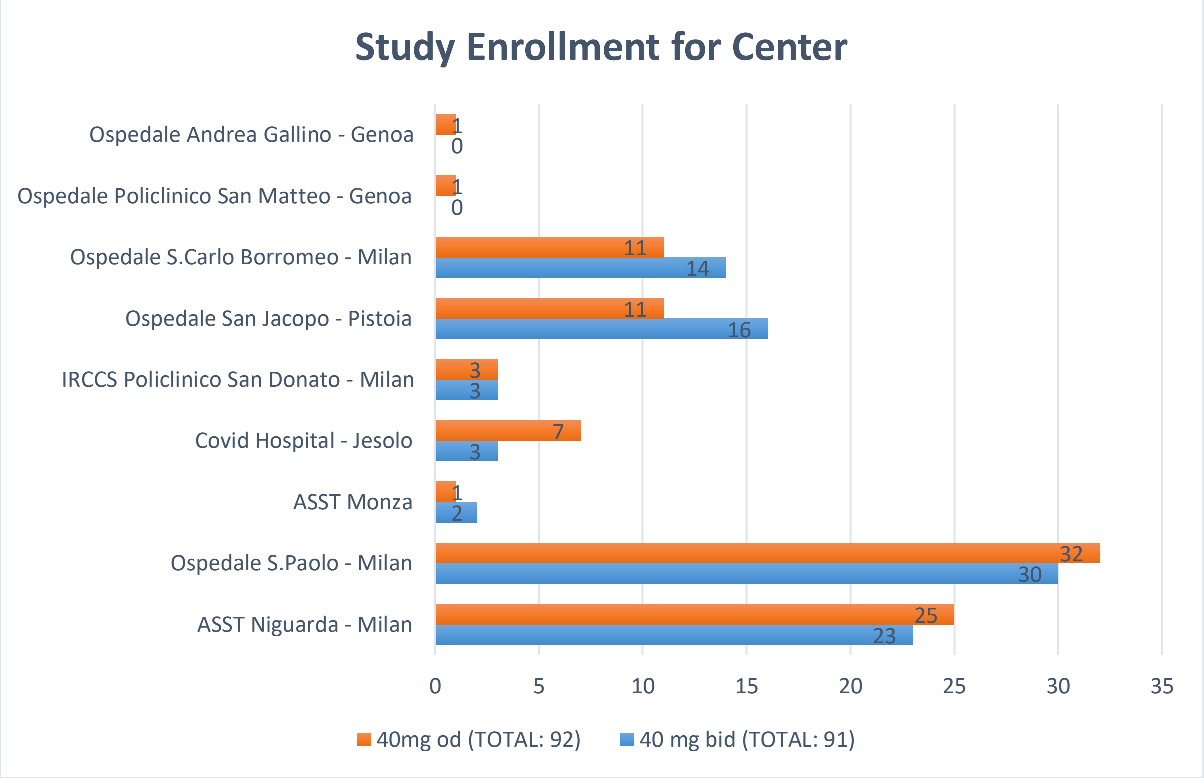
